# Longitudinal analyses reveal age-specific immune correlates of COVID-19 severity

**DOI:** 10.1101/2021.01.25.21250189

**Authors:** Sloan A. Lewis, Suhas Sureshchandra, Michael Z. Zulu, Brianna Doratt, Amanda Pinski, Micaila Curtis, Allen Jankeel, Izabela Ibraim, Nicholas Rhoades, Xiwen Jiang, Delia Tifrea, Frank Zaldivar, Weining Shen, Robert Edwards, Daniel Chow, Dan Cooper, Alpesh Amin, Ilhem Messaoudi

**Author notes:** Corresponding Author: Ilhem Messaoudi, Molecular Biology and Biochemistry University of California Irvine, 2400 Biological Sciences III Irvine, CA 92697. Equal contributions (Sloan A. Lewis, Suhas Sureshchandra).

## Abstract

Severe COVID-19 disproportionately impacts older individuals and those with comorbidities. It is estimated that approximately 80% of COVID-19 deaths are observed among individuals >65 years of age. However, the immunological underpinnings of severe COVID-19 in the aged have yet to be defined. This study captures the longitudinal immune response to SARS-CoV-2 infection in a cohort of young and aged patients with varying disease severity. Phenotypic transcriptional and functional examination of the peripheral mononuclear cells revealed age-, time, and disease severity-specific adaptations. Gene expression signatures within memory B cells suggest qualitative differences in the antibody responses in aged patients with severe disease. Examination of T cells showed profound lymphopenia, that worsened over time and correlated with lower levels of plasma cytokines important for T cell survival in aged patients with severe disease. Single cell RNA sequencing revealed augmented signatures of activation, exhaustion, cytotoxicity, and type-I interferon signaling in memory T cells and NK cells. Although hallmarks of a cytokine storm were evident in both groups, older individuals exhibited elevated levels of chemokines that mobilize inflammatory myeloid cells, notably in those who succumbed to disease. Correspondingly, we observed a re-distribution of DC and monocytes with severe disease that was accompanied by a rewiring towards a more regulatory phenotype. Several of these critical changes, such as the reduction of surface HLA-DR on myeloid cells, were reversed in young but not aged patients over time. In summary, the data presented here provide novel insights into the impact of aging on the host response to SARS-CoV2 infection.

## INTRODUCTION

Coronavirus disease-2019 (COVID-19), caused by the respiratory virus severe acute respiratory syndrome coronavirus (SARS-CoV-2), has resulted in a global pandemic with over 16 million cases and 300,000 deaths in the U.S. alone. SARS-CoV-2 targets lung airway and alveolar epithelial cells, vascular endothelial cells, and macrophages in the lung. Infection triggers a vigorous innate and adaptive immune response that culminates in viral clearance or, in the case of severe infections, the development of cytokine storm, acute respiratory distress syndrome (ARDS), and multi-organ failure (1). The estimated mortality rate of COVID-19 in the US is 1-2%, while 40-80% of patients are thought to be asymptomatic or experience mild disease symptoms (2, 3). There are numerous questions that are yet to be answered regarding the large discrepancies in disease outcomes, however, it is clear that age is a significant risk factor for severe COVID-19 and death (4-6). Indeed, infection fatality rate worldwide has been shown to increase progressively with age (0.4% at age 55, 1.4% at age 65, 4.6% at age 75, and 15% at age 85) (7). The highest proportion of patients that develop severe respiratory complications such as respiratory failure, dyspnea, pneumonia, ARDS, and death are the elderly (8-13). Aging is associated with immunosenescence, resulting in marked increase in susceptibility to respiratory viral infections, attenuated vaccine response, and increased baseline inflammation, termed inflammaging (14, 15). It is associated with decreases in naive T and B cells, increases in effector memory cells, as well as increased circulating inflammatory mediators including IL-6 and C-reactive protein (16). While there are clear age-associated immune adaptations in the lung that could lead to a reduced ability to fight SARS-CoV-2 infection (17), age-associated frailty in blood immune cells could explain paralyzed adaptive responses and poor resolution of inflammatory response in aged patients with severe COVID-19 (18). However, the specific immune mechanisms associated with increased severity of COVID-19 amongst the elderly and the immune correlates of mild and severe disease in the aged population remains elusive.

There is growing evidence that severe COVID-19 is mediated by dysregulated innate and adaptive immune responses. Elevated levels of interleukin (IL)-1, IL-6, IL-8, and CXCL10 in the blood have been associated with more severe infection or death (19-23). Furthermore, numerous studies that have profiled the peripheral immune landscape during SARS-CoV-2 infection have identified disruptions in distribution, activation as well as gene expression by single cell RNA Sequencing of immune populations. These studies have reported complex immune dysregulation in severe COVID-19, notably a pronounced decrease of HLA-DR on monocytes (21, 24, 25), an increase in non-classical CD16+ monocytes (21), and a decrease in the frequency of total dendritic cells (DCs) (25) compared to patients with moderate/mild COVID-19 and healthy controls. Additionally, it has been shown that mild disease results in increased IFN-signaling gene expression, whereas severe disease leads to impaired type-I interferon responses (26) and reduced ability to respond to a secondary stimulation in monocytes (27). Studies examining NK cells have reported reduced numbers of total NK cells with disease severity, but no differences within subsets (28-30). Similarly, NK cells have been shown to upregulate interferon response genes but have an exhausted phenotype and reduced cytolytic activity (28, 29). Another hallmark of severe COVID-19 is lymphopenia (31) that is accompanied by increased T cell activation and exhaustion (29, 32-35).

Data from these studies have begun to paint a picture of significant immune dysregulation with COVID-19 that are potentially exacerbated in aged patients (36, 37). Regardless, gaps in our knowledge remain. Notably, comprehensive longitudinal studies that include both young and aged subjects with varying disease severity (from mild to fatal) as well age-matched controls and integrated transcriptional and functional analyses are needed. In this study, we carried out a longitudinal phenotypic, transcriptional and functional analysis of innate and adaptive immune cells from young and aged COVID-19 patients compared to age-matched healthy controls. We identify age-, outcome-, and disease severity-specific alterations in the immune response to SARS-CoV-2 beginning with the discovery of differential levels of circulating immune mediators associated with age or survival. While we found comparable levels of antibodies in the young and aged patients, changes in B cell and plasmablasts frequency and gene expression with severe disease point to functional differences in humoral response. Analysis of the T cell subsets revealed heightened activation and exhaustion states with severe disease and an age-associated exacerbation of interferon and cytotoxicity signaling. Functionally, this led to immune paralysis in the T cell response. Similarly, the NK cell subsets showed discrepancies between mild and severe infection states where strong interferon induction dominated the mild response, but increased exhaustion and cytotoxicity markers were associated with more severe disease. Finally, severe COVID-19 resulted in a functional rewiring of monocytes and DC towards a more regulatory phenotype where NF-κB mediated responses were diminished. Broadly, a robust interferon gene signature was detected in individuals with mild disease but appeared late in those with severe disease. The findings presented here are critical and necessary for our understanding of anti-viral responses in the elderly and necessary for our ongoing fight against the COVID-19 pandemic.

## MATERIALS AND METHODS

### Ethics Statement

This study was approved by University of California Irvine Institutional Review Boards. Informed consent was obtained from all enrolled subjects.

### Study Participants and Experimental Design

Blood samples from patients admitted to University of California Irvine Medical Center (UCIMC) and participating in the NIH ACT-1 trial were used in these studies. Participants gave written consent to have the remainder of their blood samples used for ancillary studies related to COVID-19. Samples were stratified by disease severity - healthy donors (HD), mild/moderate COVID-19, and severe COVID-19 and age (<60 categorized as young and ≥ 60 categorized as aged). Healthy Donors (n=49; 37 young and 12 aged) included samples obtained from previous unrelated studies collected prior to 2018 and blood from seronegative healthcare workers collected after March 2020. Individuals with asymptomatic/mild disease (n=20; 10 young and 10 aged) were identified as those that tested positive for SARS-CoV-2 during their visit to UCIMC for reasons unrelated to COVID-19 symptoms (e.g. heart attack, exacerbation of auto-immune disease, elective surgeries). These samples were obtained through the COVID-19 biospecimen bank at UCIMC. A total of 47 patients with severe COVID-19 (35 young and 12 aged) were profiled, including patients with severe illness requiring hospitalization (ward) and patients with critical illness requiring intensive care unit admission without/with intubation (ICU; ICU/I). For patients with severe disease, blood was collected longitudinally over several days post symptom (DPS) onset, with median DPS being 10 and 11 days for young and aged group respectively. Six patients (2 young and 4 aged) from our analysis succumbed to disease. Due to limited sample availability, only a subset of samples were utilized in each of the assays. Detailed characteristics of the cohorts and experimental breakdown by samples is provided in Supp Table 1.

### Plasma and Peripheral Blood Mononuclear Cells (PBMC) Isolation

Whole blood samples were collected in EDTA vacutainer tubes. Peripheral blood mononuclear cells (PBMC) and blood plasma samples were isolated after whole blood centrifugation 1200 g for 10 minutes at room temperature in SepMate tubes (STEMCELL Technologies). Plasma was stored at −80^0^C until analysis. PBMC were cryo-preserved using 10% DMSO/FBS and Mr. Frosty Freezing containers (Thermo Fisher Scientific) at −80C then transferred to a cryogenic unit until analysis.

### Luminex

Immune mediators were measured using the Human XL Cytokine Discovery Premixed Kit (R&D Systems) that includes cytokines (IFNα, IFNβ, IFNγ, IL-1β, IL-10, IL-12p70, IL-13, IL-15, IL-17A, IL17E, IL-1RA, IL-2, IL-7, TNFα, TRAIL, IL-33), chemokines (CCL2/MCP-1, CCL3/MIP-1α, CCL4/MIP-1β, CCL20/MIP-3α, CCL5/RANTES, CCL11/Eotaxin, CXCL1/GROα, CXCL2/GROβ, CXCL10/IP-10, CX3CL1/Fractalkine), growth factors (GM-CSF, G-CSF, EGF, VEGF, PDGF-AA, TGFα) and effector molecules (Granzyme B, PD-L1, CD40L, Flt-3L). Samples were diluted per manufacturer’s instructions and run in duplicates on the Magpix Instrument (Luminex, Austin, TX). Data were fit using a 5P-logistic regression on xPONENT software (version 7.0c).

### Antibody ELISA

Clear 96 well, high-binding polystyrene ELISA plates were coated with 100 uL/well of 500 ng/mL SARS-CoV-2 Spike-protein Receptor-Binding Domain (RBD) or 1 ug/mL SARS-CoV-2 Nucleocapsid Protein (NP) (GenScrip) in PBS overnight at 4C. Plates were brought to RT, unbound antigen removed by flicking and blocked by adding 200 uL/well of blocking buffer for 1h at RT. Heat inactivated plasma (1:50 in blocking buffer) was added to the well and incubated for 90 minutes at RT. Responses were visualized by adding HRP-anti-human IgG (BD Pharmingen) or IgM (Cell Sciences) to the wells and incubated for another 90 min at RT. 100 uL of o-Phenylenediamine dihydrochloride (ThermoFisher Scientific) diluted with H2O2 was added as substrate. Approximately 10-20 min later, reaction was stopped with 50 ul/well of 1M HCl. ODs were read at 490 nm on a Victor3 ™ Multilabel plate reader (Perkin Elmer). Batch differences were minimized by normalizing to positive control samples run on each plate.

### PBMC and monocyte immunophenotyping

Frozen PBMCs were thawed, washed in FACS buffer (2% FBS, 1mM EDTA in PBS) and counted on TC20 (Biorad) before surface staining using two independent flow panels. For the innate panel, the following antibodies were used: CD3 (SP34, BD Pharmingen) and CD20 (2H7, BioLegend) for the exclusion of T & B lymphocytes, respectively. We further stained for CD56 (BV711, BioLegend), CD57 (HNK-1, BioLegend), KLRG1 (SA231A2, BioLegend) CD16 (3G8, BioLegend), CD14 (M5E2, BioLegend), HLA-DR (L243, BioLegend), CD11c (3.9, ThermoFisher Scientific), CD123 (6H6, BioLegend) and CD86 (IT2.2, BioLegend). For the adaptive panel, the following antibodies were used: CD4 (OKT4, BioLegend), CD8b (2ST8.5H7, Beckman Coulter), CD45RA (HI100, TONBO Biosciences), CCR7 (G043H7, BD Biosciences), CD19 (HIB19, BioLegend), IgD (IA6-2, BioLegend), CD27 (M-T271, BioLegend), KLRG1 (SA231A2, BioLegend) and PD-1 (Eh12.2h7, BioLegend). Cells were surface stained at 4C for 30 minutes, permeabilized, fixed and stained intracellularly (Ki67 (B56) for adaptive panel and Granzyme B (QA16A02) for innate panel), using using the Foxp3/Transcription Factor Staining Buffer Kit (TONBO Biosciences) per manufacturer’s instructions. Dead cells were excluded using the Ghost Dye viability dye (TONBO biosciences). Monocytes from a subset of samples were phenotyped using an additional panel. Briefly cells were thawed, incubated with Fc blocking reagent (Human TruStain FcX, BioLegend) for 10 minutes at RT, and stained at 4C for 30 minutes using the following panel: CD14 (M5E2, BioLegend), HLA-DR (L243, BioLegend), CD62L (DREG-56, BioLegend), CD163 (GHI/61, BioLegend), CCR5 (J418F1, BioLegend), CD40 (5C3, BioLegend), CCR2 (FAB151C, R&D Systems), CD64 (10.1, BioLegend), CX3CR1 (2A9-1, BioLegend), CD80 (2D10, BioLegend), CD11b (ICRF44, BioLegend), and Monocyte Blocker (Tru-Stain Monocyte Blocker, BioLegend). Samples acquired on the Attune NxT acoustic focusing cytometer (Life Technologies). Data were analyzed using FlowJo v10 (TreeStar, Ashland, OR, USA).

### Monocyte stimulation assay

Approximately 1×10e6 PBMC were cultured in the presence of RPMI (controls), bacterial and viral agonists at 37C. Bacterial agonist cocktail consists of a combination of 2ug/mL Pam3CSK4 (TLR1/2 agonist, InvivoGen), 1 ug/mL FSL-1 (TLR2/6 agonist, Sigma Aldrich), and 1 ug/mL LPS (TLR4 agonist from E. coli O111:B4, InvivoGen). Viral agonist cocktail consists of 5 ug/mL Imiquimod (R848, TLR7 agonist, InvivoGen), 1 ug/mL ssRNA (ssRNA40 LyoVec, TLR8 agonist, InvivoGen), and 5 ug/mL ODN2216 (CpG ODN, TLR9 agonist, InvivoGen). Samples were stimulated for 1 hour after which protein transport inhibitor (Brefeldin A) was added and incubated for an additional 7 hours at 37C. Cells were then washed twice in FACS buffer and surface stained using the following antibody cocktail - CD14 (M5E2, BioLegend), HLA-DR (L243, BioLegend), CD11b (ICRF44, BioLegend) for 30 minutes at 4C. Stained cells were then fixed and permeabilized using Fixation buffer (BioLegend) and incubated overnight with a cocktail of intracellular antibodies - IL-6 (MQ2-6A3, BioLegend), TNFα (MAb11, eBioScience), IFNα (LT27:295, Miltenyi Biotec). Samples were acquired on the Attune NxT acoustic focusing cytometer (Life Technologies). Data were analyzed using FlowJo v10 (TreeStar, Ashland, OR, USA).

### T cell stimulation assay

Approximately 2×10e5 PBMC were stimulated with 1 ug of each of the SARS-CoV-2 peptide pools (15-mer with 9-mer overlap) - pool 1 (Nsp1-2/Nsp4-5), pool 2 (Nsp3), pool 3 (Nsp6-12), Pool 4 (Nsp13,Nsp15-16), Pool 5 (S protein), Pool 6 (Nsp14/E/M/ORF3a/6/7a/7b/8/10) or anti CD3 (positive control) in 96 well plates for 16h at 37C and 5% CO_2_. Plates were spun, and IFN***γ*** in supernatants measured using IFN gamma Human ProcartaPlex Simplex Kit (ThermoFisher Scientific) per manufacturer’s instructions.

### Single cell RNA library preparation

Cryopreserved PBMC from each patient (n=4/group for HD and Mild; n=4/time point for severe) were thawed, washed, and stained with 1 ug/test cell-hashing antibody (TotalSeq B0251,B0254, B0256, B0260, clones LNH-95, 2M2, BioLegend) for 30 minutes at 4C. Samples were washed three times in ice cold PBS supplemented with 2% FBS and sorted on the FACSAria Fusion (BD Biosciences) with Ghost Dye Red 710 (Tonbo Biosciences) for dead cell exclusion. Live cells were counted in triplicates on a TC20 Automated Cell Counter (BioRad) and pooled in groups of 4 based on disease severity and time point. Pooled cells were resuspended in ice cold PBS with 0.04% BSA in a final concentration of 1200 cells/uL. Single cell suspensions were then immediately loaded on the 10X Genomics Chromium Controller with a loading target of 17,600 cells. Libraries were generated using the v3.1 chemistry per manufacturer’s instructions with additional steps for the amplification of HTO barcodes and library preparation using Chromium Single Cell 3’ Feature Barcoding Library Kit (10X Genomics, Pleasanton CA). Libraries were sequenced on Illumina NovaSeq with a sequencing target of 30,000 reads per cell RNA library and 2,000 reads per cell HTO barcode library.

### Single cell RNA-Seq data analysis

Raw reads were aligned and quantified using the Cell Ranger Single-Cell Software Suite with Feature Barcode addition (version 4.0, 10X Genomics) against the GRCh38 human reference genome using the STAR aligner. Downstream processing of aligned reads was performed using Seurat (version 4.0). Droplets with ambient RNA (cells fewer than 200 detected genes) and dying cells (cells with more than 25% total mitochondrial gene expression) were excluded during initial QC. Data objects from all groups were integrated using *Seurat* (version 3.2.2) (38) using the healthy, young donor samples as the reference. Data normalization and variance stabilization was performed on the integrated object using the *SCTransform* function where a regularized negative binomial regression corrected for differential effects of mitochondrial gene expression levels. The *HTODemux* function was then used to demultiplex donors and further to identify doublets, which were then removed from the analysis. Dimension reduction was performed using *RunPCA* function to obtain the first 30 principal components followed by clustering using the *FindClusters* function in Seurat. Clusters were visualized using the UMAP algorithm as implemented by Seurat’s *runUMAP* function. Cell types were assigned to individual clusters using *FindMarkers* function with a fold change cutoff of at least 0.4 and using a known catalog of well-characterized scRNA markers for human PBMC (39). Clusters from three major cell types (T and NK cells, myeloid cells, B cells) were further subsetted from the total cells and re-clustered to identify minor subsets within those groups. List of cluster specific markers identified from this study are cataloged in Supp Table 2.

### Pseudo-temporal analysis

Pseudotime trajectory of CD4 and CD8 T cells and monocytes was reconstructed using Slingshot (40). The analysis was performed on each of these subsets individually with Treg and MAIT cells excluded from T cell analysis due to unique developmental origins. For each set of cells, the UMAP dimensional reduction performed in Seurat was used as the input for Slingshot. For calculation of the lineages and pseudotime in the T cell subsets, the naive clusters were set as the root state. In the monocytes, MS1 was set as the root state.

### Differential expression analyses

Differential expression analysis was performed using MAST using default settings in Seurat. All disease comparisons were performed relative to healthy donors from corresponding age groups. Only statistically significant genes (Fold change cutoff ≥ 1.5; adjusted p-value ≤ 0.05) were included in downstream analysis.

### Module Scoring and functional enrichment

For gene scoring analysis, we compared gene signatures and pathways from KEGG (https://www.genome.jp/kegg/pathway.html) (Supp Table 3) in subpopulations using Seurat’s *AddModuleScore* function. CD8 T cell specific modules were derived from previously published work (41). NK cell exhaustion scores were calculated using aggregate expression of exhaustion markers *PDCD1* (PD-1), *LAG3, HAVCR2* (TIM-3), and *B3GAT1*. Fas-Jnk signaling scores were calculated using aggregate expression of *FAS, FASLG, DAXX, MAP3K5, MAPK8, MAPK9*, and *MAPK10*. HLA Class II module scores were calculated using aggregate expression of *HLA-DM* (*A, B*), *HLA-DP* (*A1, B1*), *HLA-DO* (*A, B*), *HLA-DQ* (*A1, A2, B1, B2*), and *HLA-DR* (*A, B1, B5*).

Over representative gene ontologies were identified using 1-way, 2-way, 4-way and 8-way enrichment of differential signatures using Metascape (42). Functional enrichment networks were edited and annotated using Cytoscape (version 3.6.1). All plots were generated using *ggplot2* and *Seurat*.

### Statistical analysis

Data sets were first tested for normality. An ordinary one-way analysis of variance (ANOVA) test was used to compare readouts from mild/severe to the healthy donors (HD) within each age group and each disease severity group between young and aged groups followed by Holm Sidak’s multiple comparisons tests. Linear regression analysis compared significant shifts in curve over horizontal line, with spearman correlation coefficient reported for each age group. Two group comparisons were tested using an unpaired t-test followed by welch’s correction. P-values less than or equal to 0.05 were considered statistically significant. Values between 0.05 and 0.1 are reported as trending patterns.

## RESULTS

### Magnitude of cytokine storm and frequency of key immune cell subsets are modulated by both age and COVID-19 severity

To comprehensively assess the peripheral immune response to SARS-CoV-2 infection with age, we performed a combination of immunological, single cell transcriptomic, and functional assays (**Figure 1A**) using blood samples obtained from 49 healthy donors (37 young and 12 aged), 20 mild/ asymptomatic COVID-19 patients (10 young and 10 aged), and 47 severely infected COVID-19 patients (35 young and 12 aged) with longitudinal sampling based on days post symptom (DPS) onset (median ∼10 DPS). Each group contained young (<60 years, median age 42.5 years) and aged patients (≥60 years, median age 69) including six patients (2 young and 4 aged) who succumbed to infection (**Supp. Table 1)**. Levels of circulating immune mediators were measured using a Luminex assay (**Supp Figure 1A**). While several soluble factors increased comparably with both mild and severe COVID-19 (IFNγ, IFNα, TRAIL, TNFα, IL-12, IL-33, IL-15, IL-13, and G-CSF), others increased to a greater extent with severe infection (MCP-1, MIP-1α, MIP-1β, CD40L, Fractalkine, GROα, GROβ, EGF, Flt-3, IFNβ, IL-1β, IL-1RA, IL-2, TGFα, and PDGF-AA) (**Supp Figure 1A**). Levels of PDL-1, Granzyme B, RANTES, and VEGF were significantly lower in aged patients with severe disease compared to their younger counterparts, while levels of IP-10, GM-CSF, IL-10, and TRAIL were significantly elevated (**Supp Figure 1A and Supp. Figure 1B**).

**Figure 1:**
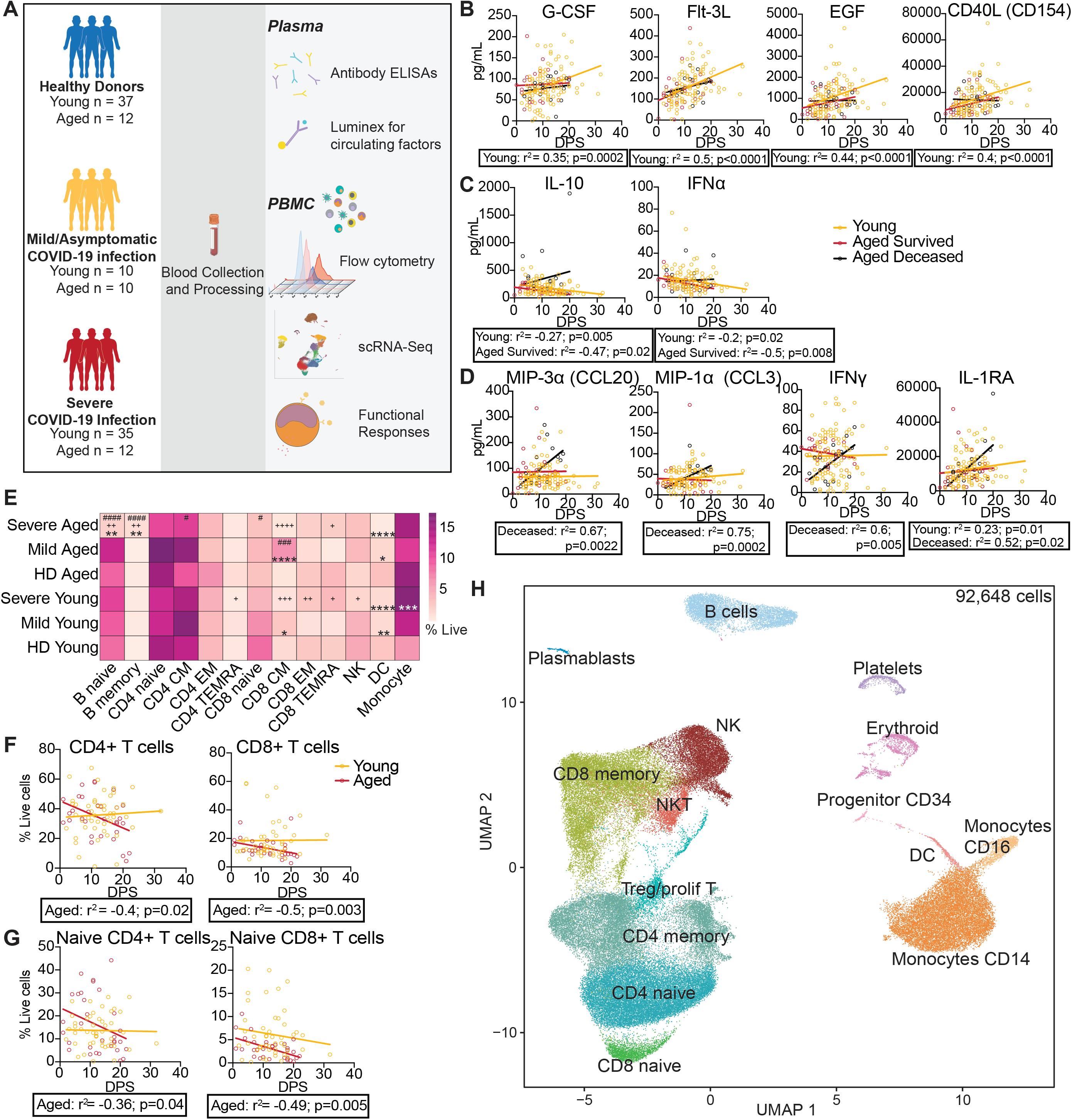
Age- and COVID-19 severity-dependent alterations in circulating factors and major immune cell subsets. (A) Experimental design for the study. Blood was collected from healthy donors (n=49; 37 young and 12 aged), mild/asymptomatic patients (n=20; 10 young and 10 aged) and longitudinally in 35 young and 12 aged patients with severe COVID-19. Immune phenotypes of PBMC and concentration of soluble mediators in plasma were determined using flow cytometry and luminex respectively. Longitudinal serological responses to SARS-CoV-2 were measured using ELISA. A subset of PBMC samples (n=4 HD, 4 Mild, 4 Severe per age group) were profiled using scRNA-Seq to determine dynamic peripheral immune adaptations of COVID-19 in mild/severe disease in young and aged subjects. (B-D) Linear regression analyses of the concentration (pg/mL) of select cytokines, chemokines, and growth factors across days post symptom onset (DPS) (E) Heatmap of proportions of major immune cell subjects derived from live singlets quantified using flow cytometry. *=significant compared to healthy donors, += significant compared to mild patients, #=significant difference between Aged Mild/Severe and Young Mild/Severe groups where */+/#=p<0.05, **=p<0.01, ***=p<0.001, ****=p<0.0001. (F-G) Linear regression analysis of frequencies of (F) total and (G) naive T cell subsets with days post symptom onset (DPS) in young and aged subjects. (H) UMAP projection of all 92,648 cells isolated from both young and aged healthy donors, patients with mild and severe (longitudinally sampled) COVID-19. Major immune cell subsets are annotated.

Linear regression analysis of soluble mediators over the course of infection in severe patients revealed that levels of G-CSF, Flt-3L, EGF, CD40L only correlated positively with days post symptom onset (DPS) in young patients (**Figure 1B**). Interestingly, levels of IL-10 and IFNα negatively correlated with DPS in patients that survived regardless of their age, but not in patients who succumbed in infection (**Figure 1C**). Contrarily, levels of MIP-3α, MIP-1α, IFNγ, and IL-1RA were significantly positively correlated with DPS only in patients that succumbed to infection (**Figure 1D**). We used supervised random forest modeling to determine factors predictive of disease severity independent of age (**Supp. Figures 1C-F**). At 1-5 DPS, IL-17A plasma levels were the strongest predictor of disease severity and positively associated with mild/moderate disease (**Supp. Figures 1C**,**D**). Later in the disease course (>10 DPS) levels of growth factors TGF-α, VEGF, and PDGF-AB/BB were the strongest predictors of disease severity and positively associated with worse outcomes (**Supp. Figures 1E**,**F**).

To assess the impact of age and COVID-19 disease severity on peripheral immune adaptations, we profiled peripheral blood mononuclear cells (PBMC) from each patient by flow cytometry. Analysis of major populations in the blood including B cells, T cells, NK cells, dendritic cells (DC), and monocytes identified significant phenotypic differences, particularly with severe disease and age (**Figure 1E**). Specifically, decreased percentages of naive B cells and a concomitant increase in memory B cell fractions was observed in aged patients with severe disease compared to healthy donors. Increased frequencies of central memory CD8 T cells (TCM) were detected in patients with mild disease, especially the aged, while no changes were detected in the CD4 T cell subsets. Moreover, severe disease was associated with a linear drop in T cell frequencies across DPS in aged patients (**Figure 1F, G**). Frequencies of DC subsets decreased significantly in both aged and young patients regardless of disease severity, whereas monocytosis was observed only in young patients with severe COVID-19 (**Figure 1E**).

To further probe the peripheral immune response during COVID-19 infection, PBMC from 16 patients (Young Mild=4; Young Severe=4; Aged Mild=4; Aged Severe=4) and 8 healthy donors (Young HD=4; Aged HD=4) were profiled by scRNA-Seq using the 10X Genomics platform. For patients with severe disease, we profiled PBMC at three time points over the course of acute infection (**Supp. Figure 2A**). An average of 2,082 cells were sequenced per patient and time point for a total of 92,648 cells (**Supp. Figure 2B**). Dimensional reduction using Uniform Manifold Approximation and Projection (UMAP) revealed distinct clustering into the major peripheral immune subsets (**Figure 1H**). Clusters were annotated based on highly expressed marker genes using Seurat’s *FindMarkers* function (**Supp. Figure 2C and Supp. Table 2**). While we did not identify significant changes in the frequency of major populations in this subset of samples, we were able to capture shifts in minor populations including plasmablasts, NKT cells, and DC with age and disease severity (**Supp. Figure 2D**).

**Figure 2:**
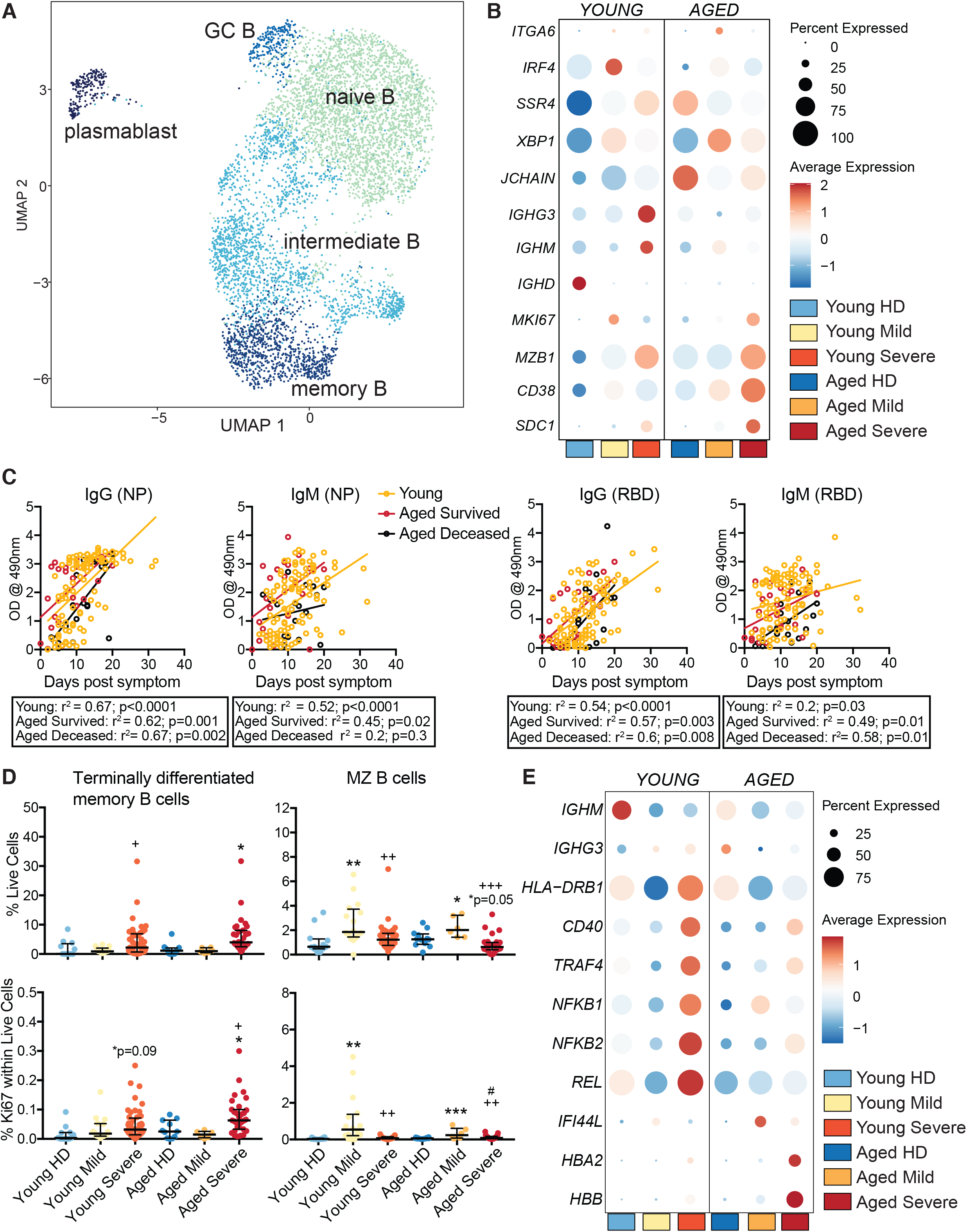
Dynamic changes in B cell and plasmablasts frequency and gene expression with age and COVID-19 severity. (A) UMAP projection of 6,334 B cells with major subsets annotated. (B) Bubble plot showing expression of select genes in the plasmablast subset. The color of the dot represents the average expression and the size represents the percentage of cells expressing that gene. (C) Linear regression analysis of antibody measurements in patients with severe COVID-19 over days post symptom onset (DPS). (D) Dot plots showing frequency of total (top) and Ki67 expressing (bottom) terminally differentiated memory B cells (left) and marginal-zone like (MZ) B cells (right) determined from flow cytometry. All percentages are reflective of fraction of total live cells. (E) Bubble plot showing expression of select genes in memory B cell subsets. The color of the dot represents the average expression and the size represents the percentage of cells expressing that gene. *=significant compared to healthy donors, += significant compared to mild patients, #=significant difference between Aged Mild/Severe and Young Mild/Severe groups where */+/#=p<0.05, **=p<0.01, ***=p<0.001, ****=p<0.0001.

### Reduced activation of memory B cell subsets in aged patients with severe disease but no differences in antibody levels

To assess the impact of disease severity and age on humoral responses to SARS-CoV-2 infection, we reclustered the B cells and plasmablasts and identified five subsets based on expression of key B cell markers (**Figure 2A and Supp. Figure 3A**). Modest increases in proportions of plasmablasts were observed with disease but were more prominent in younger patients with mild disease (**Supp. Figure 2D and Supp. Figure B**). Differential expression analysis within each group relative to age-matched healthy donors revealed increased expression of plasma cell genes *MZB1, CD38*, and *SDC1* in severe disease in plasmablasts regardless of age while expression of immunoglobulin heavy chain genes *IGHG3* and *IGHM* was only increased in young patients with severe disease (**Figure 2B**). Mild disease was associated with upregulation of genes associated with B cell commitment to plasma cells and antibody production (*IRF4, XBP1*). Interestingly, expression of *SSR4*, important for B cell effector function, was increased with infection in young patients but decreased in aged patients (**Figure 2B**).

**Figure 3:**
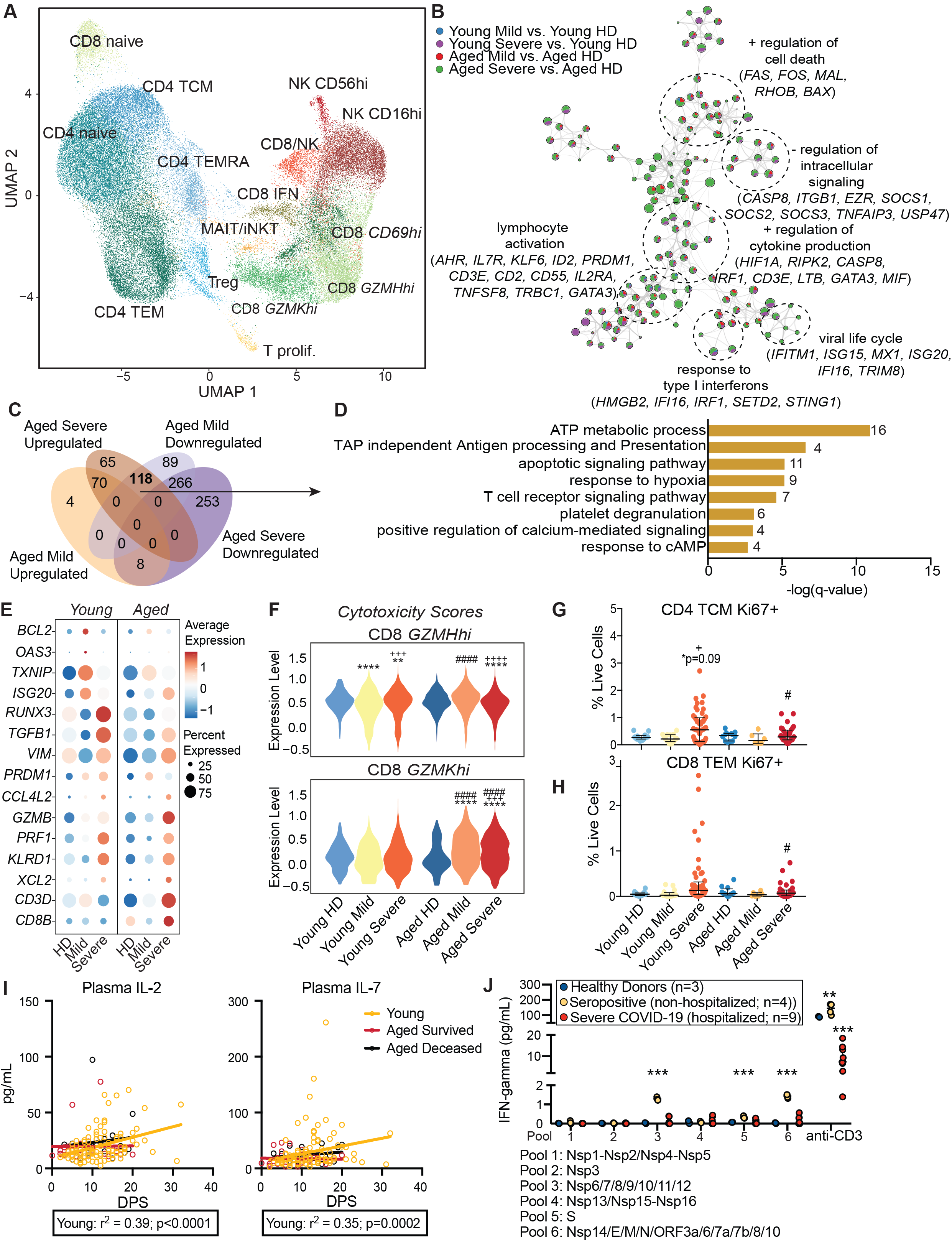
Heightened activation and cytotoxicity of T cells from aged patients with severe COVID-19. (A) UMAP projection of 66,656 lymphocytes reclustered from the main UMAP to identify T and Natural Killer (NK) cell subpopulations at a higher resolution. Major subsets of CD4 T cells, CD8 T cells, and NK cells are highlighted. (B) Network of functional enrichment of genes up-regulated with COVID-19 in CD4 TEM T cell cluster. Themes of highly related GO terms are grouped within dashed circles. GO terms are shown as pie charts that depict the relative DEG contributions from the four groups being compared. (C) Venn diagram of DEG detected in CD4 TEMRA clusters in aged subjects with mild and severe COVID-19. (D) Bar graph showing GO enrichment of DEGs up-regulated with severe COVID-19 but down-regulated in mild COVID-19 aged subjects within the CD4 TEMRA cluster. Gene numbers associated with each term are indicated next to each term. Size of the bar indicates the significance of the enrichment. (E) Bubble plot of a subset of genes that are differentially responsive in mild versus severe COVID-19 within memory CD8 T cell clusters. (F) Violin plots comparing cytotoxicity module scores within *GZMH*^hi^ and *GZMK*^hi^ CD8 T cell clusters. (G-H) Dot plots comparing frequencies of (G) proliferating CD4 TCM and (H) proliferating CD8 TEM with mild and severe COVID-19 in young and aged subjects measured using flow cytometry. *=significant compared to healthy donors, += significant compared to mild patients, #=significant difference between Aged Mild/Severe and Young Mild/Severe groups where */+/#=p<0.05, **=p<0.01, ***=p<0.001, ****=p<0.0001. (I) Linear regression of plasma IL-2 and IL-7 levels and days post symptoms (DPS) within young (mustard) and aged subjects (survived - red; deceased - black) with severe COVID-19. (J) Secreted IFN***γ*** levels following *ex vivo* PBMC stimulation with either SARS-CoV-s peptide pools or anti-CD3 in seronegative, seropositive convalescent health care workers, and severe COVID-19 patients. Differences in secreted IFN***γ*** was tested using one-way ANOVA followed by Holm-Sidak’s multiple test comparison relative to seronegative healthy donors.

Despite these gene expression differences, levels of IgG and IgM specific to nucleocapsid protein (NP) and receptor-binding domain (RBD) were comparable in young and aged patients (**Supp. Figure 3C**). Independent of age, antibodies against NP showed a stronger positive correlation with DPS compared to antibodies against RBD (**Figure 2C**). Poor induction of NP-specific IgM levels was detected in patients who succumbed to COVID-19. Additionally, levels of NP- and RBD-specific IgG were lower in aged patients who succumbed to disease during DPS 1-5 (**Supp. Figure 3D**).

We next identified disease and age-specific changes in frequency and gene expression within memory B cell subsets. Independent of age, frequency of total and proliferating terminally differentiated B cells was increased in patients with severe disease, while that of total and proliferating marginal zone-like (MZ) B cells was increased only in patients with mild disease (**Figure 2D**). However, while severe disease was associated with induction of genes associated with B cell activation (*CD40, TRAF4, NFKB1/2* and *REL*), these changes were less pronounced in aged subjects (**Figure 2E**). Interestingly, expression of heavy chain gene *IGHG3* increased with infection in young patients but decreased in aged patients. Finally, *IFI44L* was increased to a greater extent with mild disease than severe disease (**Figure 2E**).

### Severe COVID-19 in aged patients is associated with heightened activation and apoptotic signaling in CD4 T cells

Given the reduction in the frequencies of naive T cells in aged subjects with COVID-19 **(Figures 1F, G)**, we next investigated the impact of age and COVID-19 severity on phenotypic and transcriptional changes in memory T cell subsets. T cells and NK cells were reclustered from the original UMAP **(Figure 1H)** to identify subsets at a higher resolution. Using the *FindMarkers* function in Seurat, we identified naive CD4 T cells (expressing high *CCR7, IL7R*), central memory CD4 T cells (TCM) (expressing *CCR7, LEF1, IL7R*), effector memory CD4 T cells (TEM) (expressing high *CD69, S100A4*), and CD4 TEMRA (expressing low *CCR7* but high *PTPRC* and *RORA*) **(Figure 3A and Supp. Figure 4A)**. Pseudotime analysis of CD4 T cells placed cells along a trajectory arising from naive CD4 followed by TCM CD4 and ending in TEM **(Supp. Figure 4B)**. While the frequencies of CD4 TEM did not vary significantly with mild or severe disease, significant transcriptional changes were observed with severe disease relative to healthy donors only in the aged subjects (**Figure 3B**). Specifically, up-regulation of pathways associated with lymphocyte activation (*CD3E, IL2RA, IL7R*), response to type-I interferons (*IFI6, IRF1*), and positive regulation of cell death (*FAS*) **(Figure 3B)**, were observed in T cells from aged patients with severe disease. Interestingly, genes associated with antiviral immunity (*ISG15, ISG20, MX1, TRIM8*) were up-regulated predominantly in aged patients with severe disease **(Figure 3B)**.

**Figure 4:**
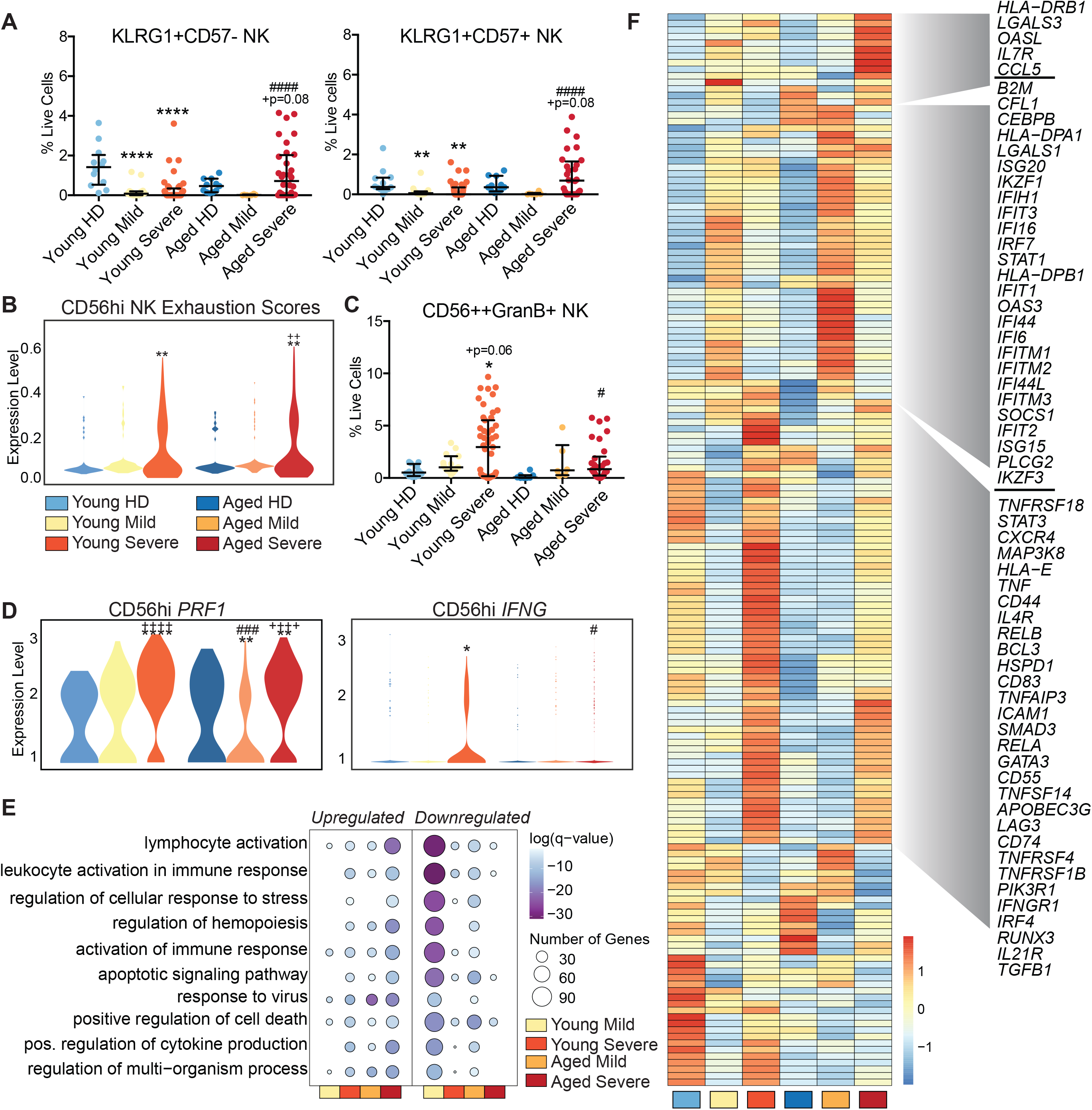
Increased exhaustion of NK cells in aged patients with severe COVID-19. (A) Dot plots comparing frequencies of KLRG1+ NK cells expressing CD57 using flow cytometry. (B) Violin plots comparing exhaustion module scores for CD56^high^ (*NCAM*^*high*^) NK cell subset. (C) Dot plots comparing granzyme B expressing CD56++ NK cells measured using flow cytometry. (D) Gene expression of cytotoxic markers *PRF1* and *IFNG* within CD56^high^ NK cells. (E) Bubble plot representing functional enrichment of DEG in CD16^high^ (*FCGR3A*^high^) NK cells from aged and young subjects with mild and severe COVID-19. Color and size of the bubble represents statistical significance and number of genes respectively. (F) Clustered heatmap comparing normalized expression of genes enriching to GO term “*lymphocyte activation*” from CD16^high^ NK subset obtained from patients with mild and severe COVID-19. Colors represent scaled gene expression ranging from blue (low) to red (high). Select genes are highlighted. *=significant compared to healthy donors, += significant compared to mild patients, #=significant difference between Aged Mild/Severe and Young Mild/Severe groups where */+/#=p<0.05, **=p<0.01, ***=p<0.001, ****=p<0.0001.

Severe COVID-19 was associated with enrichment of CD4 TEMRA subset, most notably in young patients **(Supp. Figure 4C)**. Moreover, this subset was transcriptionally dysregulated with severe disease in aged patients compared to their control counterparts **(Figure 3C)**. Specifically, expression of genes associated with ATP metabolism, calcium signaling and response to hypoxia (including cytotoxic molecules such as *GZMA, GZMB*, and *KLRD1*) was decreased in aged patients with mild disease and increased in aged patients with severe disease **(Figure 3D)**. Genes that were uniquely down-regulated with severe disease in aged patients enriched to gene ontology (GO) terms associated with antiviral immunity and type-I IFN signaling **(Supp. Figure 4D)**. A consistent theme across in both TEM and TEMRA clusters was up-regulation of genes that map to cell death signaling GO terms with severe disease, confirmed by increased scoring of Fas signaling module in memory CD4 T cells with severe COVID-19, especially aged patients **(Supp. Figure 4E and Supp. Table 3)**.

### Exacerbated IFN and cytotoxicity signaling in CD8 T cells from aged patients with severe COVID-19

Single cell analysis of CD8 T cells (expressing *CD8A*) identified 4 memory subset clusters in addition to naive CD8 T cells (expressing high *CCR7* and *LEF1*) **(Figure 3A and Supp. Figure 4A)**. These memory clusters were identified as CD8 IFN (expressing high levels of ISGs such as *IFIT2, IFIT3*), activated memory CD8 (*CD69*^high^ expressing *CCL4*, and *IFNG*), and two subsets of cytotoxic CD8 T cells (*GZMH*^high^ and *GZMK*^high^) (**Figure 3A and Supp. Figure 4A**). Pseudotime analysis placed the CD8 T cells along a trajectory starting from naive CD8, antiviral CD8, activated CD8, and terminating in cytotoxic CD8 subsets **(Supp. Figure 4F)**. The phenotypes of these subsets were confirmed using module scoring - CD8 *GZMH*^high^ and expressed the highest levels of cytotoxicity genes; CD8 *CD69*^high^ the highest cytokine score; and the CD8 IFN cluster the highest interferon module scores (**Supp. Figure 4G**). We observed a modest decrease in naive CD8 T cells in the young group with COVID-19 **(Supp. Figure 4H)** and a modest increase in CD8 IFN cluster in aged patients with mild disease **(Supp. Figure 4I)**. No differences in the frequencies of CD8 *GZMK*^high^ or *GZMH*^high^ clusters were observed with mild/severe disease**(Supp Figure 4J, K)**. However, we observed a significant reduction in CD8 *GZMK*^high^ cells in aged patients who succumbed to disease compared to those who did not **(Supp Figure 4K)**. Comparisons of COVID-19 associated differentially expressed genes in memory CD8 T cells revealed up-regulation of pro-survival (*BCL2*) and antiviral factors (*OAS3, TXNIP*) with mild disease and enrichment of factors associated with the tissue resident CD8 T cell development program (*RUNX3, VIM, TGFB1*) in severe patients **(Figure 3E)**. More importantly, while CD8 T cells from both young and aged severe COVID-19 patients expressed elevated levels of chemokines (*CCL4L2, XCL2*) and cytotoxic molecules (*PRF1, GZMB, KLRD1*) **(Figure 3E)**, the overall cytotoxicity scores in *GZMK*^high^ clusters were significantly higher in aged patients **(Figure 3F)**. As observed in the CD4 TEM cluster, severe disease was associated with signatures of heightened CD8 T cell activation (*CD3D* and *CD8A*) **(Figure 3E)**. Finally, our analyses revealed significant up-regulation of interferon signaling in all CD8 T cell clusters with mild disease in both young and aged subjects **(Supp. Figure 5A)**, being significantly higher in young mild relative to aged mild (p< 0.0001). However, severe disease in the aged was associated with elevated interferon signaling scores relative to severe young (p<0.0001), peaking at days 18-19 DPS **(Supp. Figures 5A, B)**.

**Figure 5:**
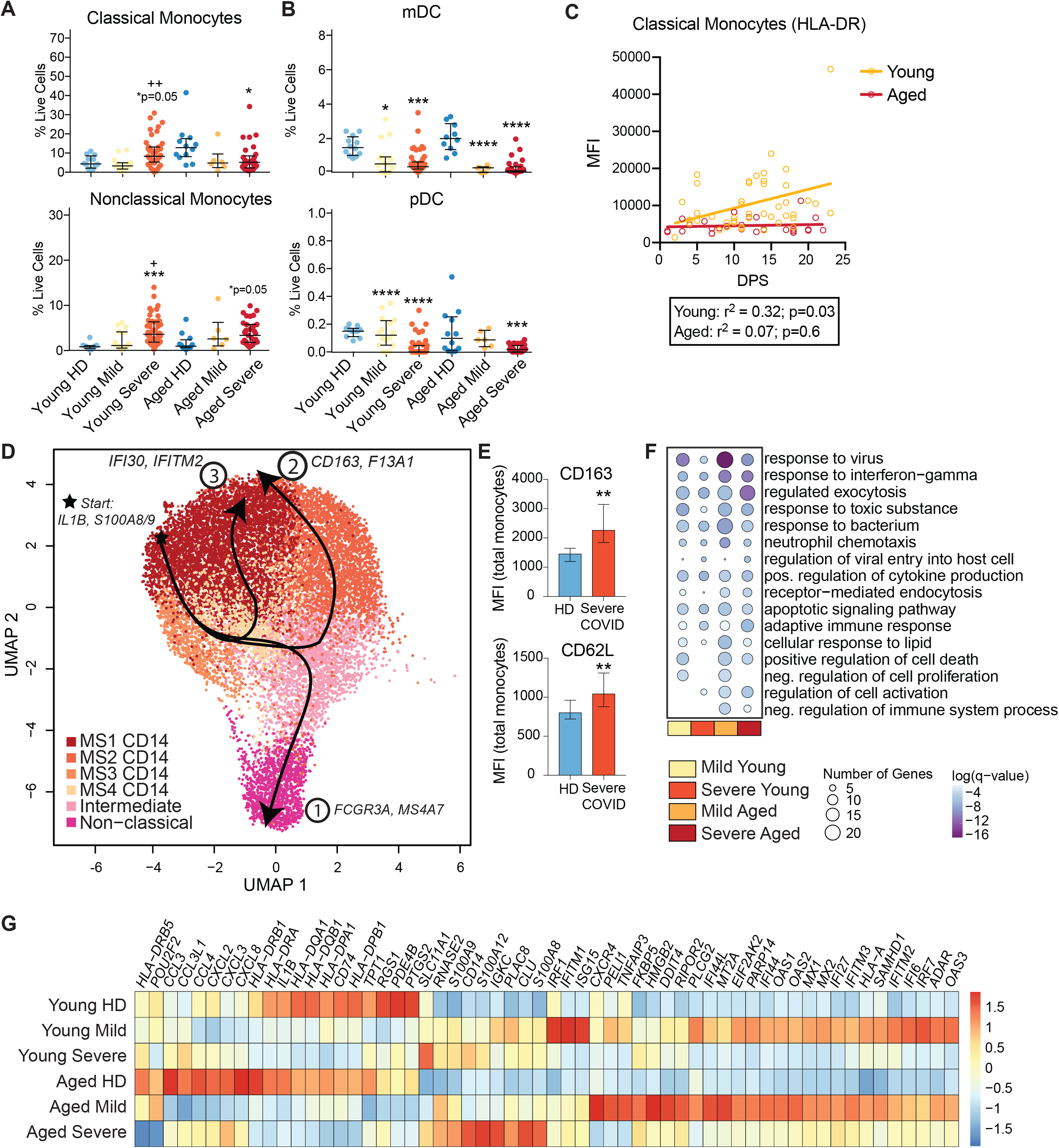
Transcriptional and functional rewiring of myeloid cells with severe COVID-19. (A-B) Dot plots comparing frequencies of (A) classical monocytes and non-classical monocytes as well as (B) myeloid DC (mDC) and plasmacytoid DC (pDC) using flow cytometry. (C) Linear regression analysis of HLA-DR surface expression on classical monocytes as a function of days post symptom (DPS) in young and aged subjects. (D) UMAP projection of 15,922 monocytes (classical and non-classical) clustered from the main UMAP. The three trajectories identified by slingshot are highlighted with markers describing each trajectory. (E) Surface expression changes in CD163 and CD62L on total monocytes from healthy donors (n=5) and patients with severe COVID-19 (n=15). (F) Bubble plot of gene ontology terms derived from COVID-19 induced DEG in classical monocytes from aged and young subjects. Color and size of the bubble represents statistical significance and number of genes respectively. (G) Clustered heatmap comparing normalized transcript levels of genes enriching to GO terms “*response to virus*” and “*response to interferon-gamma*” within classical monocytes from young and aged patients with mild and severe COVID-19. Colors represent scaled gene expression ranging from blue (low) to red (high). *=significant compared to healthy donors, += significant compared to mild patients, #=significant difference between Aged Mild/Severe and Young Mild/Severe groups where */+/#=p<0.05, **=p<0.01, ***=p<0.001, ****=p<0.0001.

### Severe COVID-19 is associated with immune paralysis in the T cell compartment

Single cell RNA-Seq analysis revealed enrichment of proliferating T cell subsets with severe COVID-19 in both young and aged patients **(Supp. Figure 5C)**. We confirmed these observations in a larger set of samples by measuring Ki67 expressing using flow cytometry. Our analysis revealed that proliferating T cells in CD4 and CD8 subsets were prominent within central memory **(Figure 3G)** and effector memory **(Figure 3H)** compartments respectively. This induction of proliferating CD4 and CD8 T cells was significantly reduced in aged severe patients compared to young counterparts (**Figure 3G, H**). In line with this defect, we observed a progressive increase in T cell maintenance factors IL-2 (r^2^=0.39, p<0.0001) and IL-7 (r^2^=0.35; p<0.001) only in young patients with severe disease **(Figure 3I)**. Furthermore, a sharp decline in IL-2 signaling signatures was detected in memory CD4 T cells from aged subjects with severe disease only (r^2^=-0.28) **(Supp. Figure 5D)**. Finally, we observed modest reduction in frequencies of both proliferating T cells and regulatory T cells in aged patients who succumbed to the disease **(Supp. Figure 5C)**.

Next, we tested if severe SARS-CoV-2 infection alters the ability of T cells to respond to SARS-CoV-2 peptides or polyclonal stimulation (n_HD_=3; convalescent=4; n_severe_=9). While T cells from convalescent patients secreted significantly higher levels of IFN***γ*** relative to healthy donors in response to several of the viral peptide pools, these responses were significantly attenuated with severe disease **(Figure 3J)**. We also noted weaker T cell responses to polyclonal stimulation in patients with severe disease relative to healthy donors (p<0.01) and an independent group of convalescent health care workers (sero-positive) (p<0.001) **(Figure 3J)**.

### Delayed induction of IFN signaling but increased exhaustion and cytotoxicity in NK cells with severe COVID-19

Given their critical role in antiviral immunity during acute infection, we next examined the impact of COVID-19 disease severity and age on NK cell phenotypes. The two major populations of NK cells were identified by scRNA-Seq (NK CD16^hi^, NK CD56^hi^) based on expression of *FCGR3A* and *NCAM1* (**Figure 3A, Supp. Figure 6A**). We also identified a cluster of cells expressing NK and CD8 T cell markers (*CD8A, PLCG2*) which we call CD8/NK T cells. The percentage of CD8/NK T cells increased significantly in young patients and modestly in aged patients with severe disease, but negatively correlated with DPS (**Supp. Figure 6B, C**). No differences in major NK cell populations were detected with mild/severe disease in young or aged subjects, either by flow cytometry or scRNA-Seq (**Supp. Figure 6D)**. However, the frequency of NK cells expressing activation marker KLRG1 and/or terminal differentiation marker CD57 was decreased in young patients with severe disease but increased in aged patients with severe disease (**Figure 4A**). Additionally, NK cell exhaustion module scores from the CD56^hi^ cluster determined from the scRNA-Seq analysis were increased with severe disease (**Figure 4B**). Moreover, induction of a robust cytotoxic program with severe disease was evident from significant increase in CD56++ GranzymeB+ (flow cytometry) and expression of *PRF1* and *IFNG* (single cell RNA), which were less pronounced in severe aged (**Figure 4C, D**).

**Figure 6:**
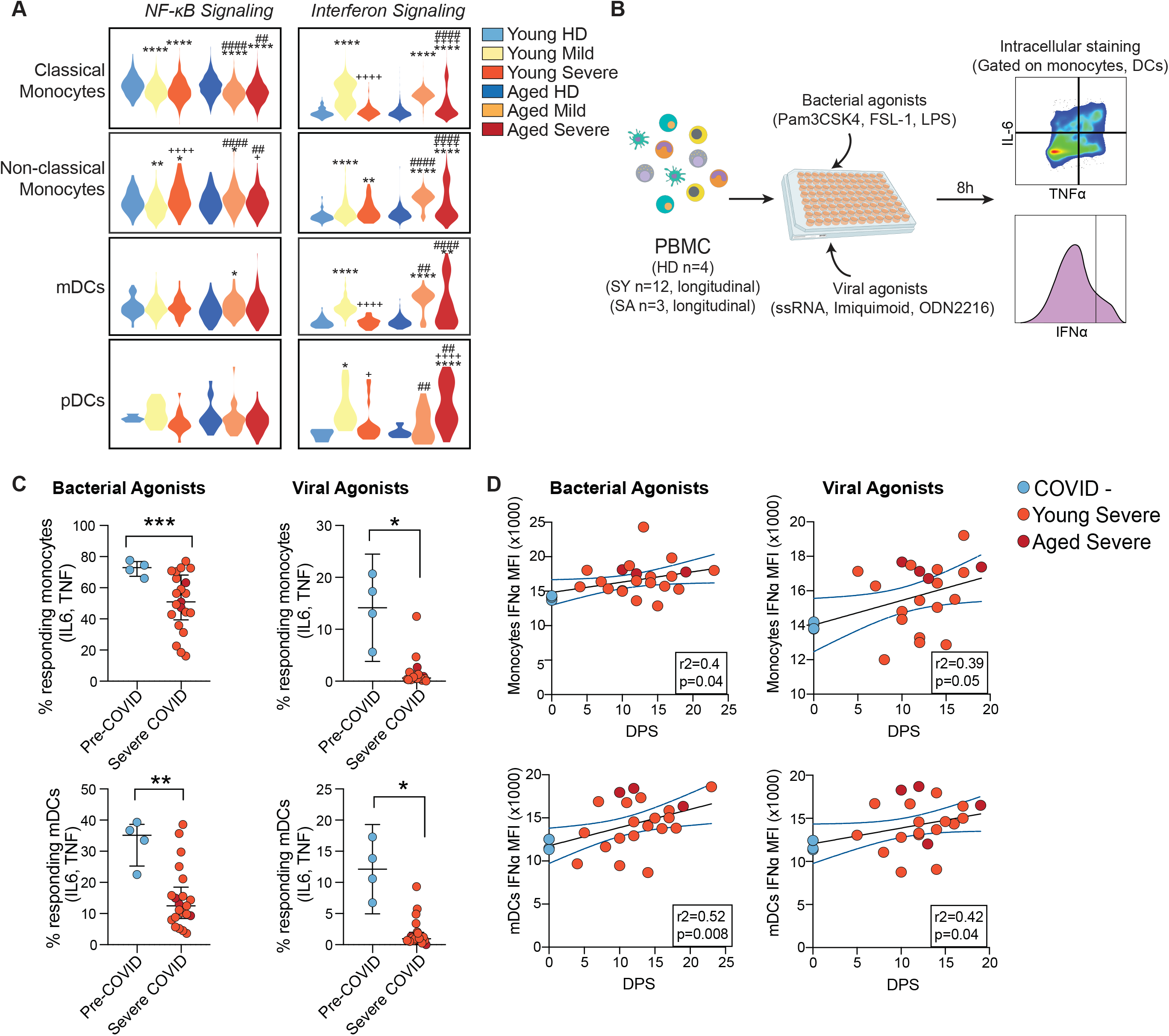
Dysregulation of Toll-like receptor responses in myeloid cells from patients with severe COVID-19. (A) Violin plots comparing interferon and NF-κB signaling module scores in monocyte and DC subsets from mild and severe COVID-19. (B) Experimental design to measure functional responses of monocytes and myeloid DCs from patients with severe COVID-19. PBMC collected longitudinally over the course of infection from patients and healthy donors were stimulated with bacterial agonists (Pam3CSK4, LPS, and FSL-1) and viral agonists (ssRNA, Imiquimod, ODN2216) separately for 8 hours. Cytokine responses (TNFα, IL-6, and IFNα) were measured using intracellular staining followed by flow cytometry. (C) Dot plots comparing frequencies of monocytes (CD14+, top) and myeloid DCs (HLA-DR+CD11c+, bottom) producing IL-6, TNFα, or both, following *ex vivo* stimulation with bacterial (left) and viral agonists (right). (D) Scatter plots with linear regression lines tracking mean fluorescence of IFNα in IFNα+ monocytes (top) and IFNα+ myeloid DCs (bottom) following stimulation with bacterial (left) and viral agonists (right). X-axis denotes days post symptom onset.

Even though frequencies of CD16++ NK cells did not change with disease severity/age (**Supp. Figure 6D**), we observed significant changes in their transcriptional program with COVID-19. DEG from each disease group compared to their age-matched healthy donors enriched to GO processes associated with leukocyte activation, cell death, and response to virus (**Figure 4E**). Interestingly, the same pathways that were being upregulated with severe disease in the aged patients including *lymphocyte activation* and *leukocyte activation in immune response* were strongly downregulated with mild infection in the young patients. Indeed, genes involved in cell migration (*LGALS3* and *CCL5*) and activation (*HLA-DRB1*) were highly upregulated with severe disease in aged patients (**Figure 4F**). On the other hand, antiviral signatures including numerous interferon stimulated genes (ex. *ISG20, IFIT3, IFITM1*) as well as *B2M, IRF7*, and *STAT1* were upregulated with mild disease (**Figure 4F**). Finally, a number of molecules associated with cytokine signaling (*TNFSF4, TNFRSF18, STAT3, TNF, MAP3K8, RELA*) and a number of cytokines and chemokines and their receptors (*CXCR4, IFNGR1, IL21R, IL4R*) were upregulated with severe disease but to a less extent in aged patients (**Figure 4F**). This trend was confirmed from functional module scoring which showed dampened cytokine and chemokine receptor signaling scores with severe disease in the aged (p=0.09) (**Supp. Figure 6E**). Module scoring revealed increased expression of interferon signatures in both NK subsets with mild disease, but exacerbation of this signal with age in both CD56^hi^ (p<0.05) and CD16^hi^ (p<0.0001) subsets **(Supp. Figure 6F)**. Further examination of the interferon response module in the severe patients across DPS showed this increase occurred only at later infection timepoints irrespective of age (**Supp. Figure 6G**).

### Rewiring of monocytes and DCs towards a regulatory phenotype and reduced NF-κB mediated responses with severe COVID-19

Our initial PBMC analysis showed increased monocyte frequency in young patients only (**Figure 1E**). However, within monocyte subsets, we detected increases in non-classical monocytes in both young and aged subjects while frequencies of classical monocytes were significantly reduced in the aged patients with severe disease (**Figure 5A**). In contrast, proportions of both DC subsets - pDCs and mDCs was reduced in both young and aged COVID-19 patients regardless of disease severity (**Figure 1E and Figure 5B**). Additionally, expression of MHC Class II molecule HLA-DR and activation marker CD86 were broadly decreased with COVID-19, although the decrease was more significant with severe disease (**Supp. Figure 7A, B**). Regression analysis of HLA-DR expression revealed a progressive recovery in its expression on classical monocytes over the course of infection in young but not in aged subjects (**Figure 5C**).

Reclustering of the myeloid cell subsets from the scRNA-Seq data identified four clusters of classical monocytes (MS1-MS4), intermediate and non-classical monocytes, as well as mDCs and pDCs based on the expression of canonical markers (*CD14, FCGR3A, FCER1A, LILRA4*) (**Figure 5D and Supp. Figure 7C, D**). Scores of the MHC Class II module were significantly reduced in classical (mild and severe disease) and non-classical (severe disease) monocytes in both age groups (**Supp. Figure 7E**). However, severe age was associated with further reduction in MHC Class II module scores in classical monocytes (**Supp. Figure 7E)**. We also detected minor redistribution of monocyte subsets with disease severity and age. Specifically, the MS1 cluster (expressing high levels of *CXCL8* and *IL1B*) (**Supp. Figure 7D**) decreased in young patients, while the MS4 cluster (expressing antiviral genes) (**Supp. Figure 7D**) increased in patients with mild disease (**Supp. Figure 7F, G**). Trajectory analysis revealed three unique lineages within the monocyte subsets (**Figure 5D**). Lineage 1 started in the MS1 cluster and ended with the non-classical monocytes. Lineage 3 moved through the viral cluster, upregulating *IFI30* and *IFITM2*, before returning back to MS1. Interestingly, lineage 2 which moved from MS1 towards MS2, was driven almost exclusively by cells from aged patients, and was defined by increased expression of regulatory markers *CD163* and *F13A1* (**Figure 5D**). Skewing of monocytes towards a regulatory phenotype with severe disease was confirmed by increased surface expression of CD163 as well as adhesion molecule CD62L on monocytes from patients with severe COVID-19 (**Figure 5E**). Additionally, surface expression of CX3CR1 and CD11b was downregulated while that of CCR5 and CD64 upregulated with severe disease (**Supp. Figure 7H**). Regression analysis further indicated that expression of CCR5 and CD64 progressively decreased with disease progression (**Supp. Figure 7H**).

Differential gene expression analysis within classical monocytes revealed significant enrichment to GO terms *response to virus* and *response to IFNγ* pathways in all patients, that was most significant in monocytes from aged subjects with mild disease (**Figures 5F**). A handful of ISGs were up-regulated more significantly in young patients with mild disease (*ISG15, IFITM1, IRF1*) (**Figure 5G**). However, the overall interferon signaling signature was heightened in aged patients with severe disease (**Figure 6A**). Similar trends were observed in nonclassical monocytes (**Supp. Figure 8A**). Interestingly, within classical monocytes expression of alarmin molecules (S100A8, S100A9, S100A12) was increased while that of chemokines was decreased (*CXCL8, CCL3, CCL4, CXCL2, CXCL3*) with severe disease (**Figure 5G**). Moreover, the magnitude of that change was higher in the aged (**Figure 5G**). Finally, significant changes in expression of genes associated with wounding response, myeloid cell activation, and hematopoiesis were observed in non-classical monocytes with both mild and severe disease in the aged subjects (**Supp. Figure 8A**). DEG analysis in the mDC subset revealed significantly increased expression of IFN-responsive genes *ISG20, MX1, STAT1*, and *IFI44L* with disease in aged patients (**Supp. Figure 8B)**. RIGI signaling was also increased in the mDC population from the aged patients with only a modest increase in young patients (**Supp. Figure 8C**). Finally, module scoring of all the myeloid subsets revealed a decrease in NF-κB signaling with COVID-19 (mild and severe disease in classical monocytes; mild disease in nonclassical monocytes) (**Figure 6A**).

To analyze the functional capacity of myeloid cells to respond to a secondary infection, we stimulated total PBMC from a subset of our patient cohort with either a viral or bacterial agonist cocktail and measured the production of IL-6, TNFα, and IFNα in monocytes and DC (**Figure 6B**)(n_HD_=4; n_severe_=15). In response to either bacterial or viral agonists, a lower percentage of both monocytes and DC in severe patients produced IL-6 and TNFα, regardless of age (**Figure 6C**). Stratifying the IFNα responses across DPS revealed that in severe patients, there was a positive correlation of interferon production with DPS (**Figure 6D**). These data suggest that severe COVID-19 leads to a re-wiring of the inflammatory response pathways in myeloid cells with implications for response to secondary infections.

## DISCUSSION

The clinical presentation of COVID-19 is highly heterogeneous, influenced by host factors such as age, sex, BMI, and underlying medical conditions, notably diabetes, hypertension, and cardiovascular disease. Advanced age is a major driver of severe disease with 78.2% of deaths occurring in people ≥65 years of age (43). This increased vulnerability of older individuals to respiratory viral infections is mediated by age-associated deterioration of immune function (immunosenescence) and baseline inflammation (inflammaging), which can exacerbate host response to COVID-19 (18). While recent studies have shed light on host response to mild and severe COVID-19, most of these studies used mixed cohorts; thus, the immune correlates of the disease in aged populations remain unclear. To address this question, we profiled the inflammatory environment using luminex and the immune landscape using flow cytometry and single cell RNA-sequencing in blood samples from young and aged patients with mild and severe COVID-19 and compared these measurements to those obtained in age-matched healthy donors. Our analysis revealed that independent of disease severity, COVID-19 was associated with a significant shift in plasma inflammatory factors, including elevated Th1 (IFN***γ***, TNFα, IL-12) and Th2 cytokines (IL-13, G-CSF). Severe disease, however, was associated with a more prominent elevation of pro-inflammatory cytokines (IL-1β, IL-2, IL-7) and chemokines (MCP-1, MIP-1α, MIP-1β, CX3CL1, GRO-α, and GRO-β), and growth factors (TGF-α, PDGF-AA) compared to both healthy donors and those with mild disease. These observations highlight a cytokine storm that is exacerbated with severe disease, as previously reported (21, 23). Importantly, our analysis revealed that older subjects with severe disease showed significantly higher induction of GM-CSF, IP-10, and IL-10 but lower levels of PD-L1, VEGF, and RANTES compared to their younger counterparts. Elevated GM-CSF, a myelopoietic pro-inflammatory growth factor combined with lower levels of regulatory factors PD-L1 suggests a heightened inflammatory response in aged patients with severe COVID-19 (44). Lower levels of RANTES coupled with higher levels of IP-10 indicate dysregulated recruitment of T cells with potentially greater adhesion to endothelial cells (45-47), while lower levels of VEGF suggests delayed control of tissue damage. Furthermore, linear regression analysis showed progressive increase in pro-inflammatory mediators MIP-3α, MIP-1α, and IFN***γ*** and a failure to downregulate IL-10 and IFNα in patients who succumbed to infection. Collectively, our data indicate discordant inflammatory and regulatory responses in aged subjects with severe COVID-19, particularly in non-survivors. Our analysis also indicate that lower levels of IL-17 during the first 5 DPS was associated with poor prognosis (ICU admission/intubation and death). This observation differs from recent reports that high levels of IL-17 (21) and Th17 cells (48) is correlated with immunopathology of severe COVID-19 and the proposed use of IL-17 blockade as a therapeutic avenue to limit acute lung injury (49). This discrepancy highlights the need for careful evaluation of the timing for administering immune modulators in the context of COVID-19.

We next utilized a combination of flow cytometry, single cell RNA sequencing and functional assays to better understand age-specific immunological changes with mild and severe COVID-19. As previously reported, substantial induction of plasma B cells was observed with mild disease (29, 50-52), though less prominent in aged individuals. Furthermore, gene signatures associated with commitment to plasma cells were more robust with mild disease. Interestingly, while mild disease was associated with accumulation and proliferation of marginal zone B cells, severe disease was associated with accumulation of IgD-CD27-memory B cells, which have been linked to severe outcomes following COVID-19 (50). Finally, transcriptional signatures associated with B cell activation and NF-κB signaling were dampened in aged subjects with severe COVID-19. Coupled with lower levels of plasma CD40LG in aged subjects with severe disease, these observations suggest weak T follicular helper cell responses in the aged. Surprisingly, plasma antibody levels (IgG or IgM) against SARS-CoV-2 proteins RBD and NP increased similarly with DPS in young, aged, and deceased subjects. However, our analysis did not evaluate age-related differences in avidity, neutralization, and effector functions of antibodies, which should be a focus of future studies.

T cell lymphopenia is a hallmark of acute viral infection, including severe COVID-19 (52) (24) (53) (54), especially within the CD8 T cell compartment (50, 51, 54) and with advanced age (52). This is often accompanied by a concomitant accumulation of memory T cells, particularly in EM and TEMRA subsets (53) (50) (25). Our analyses revealed a significant drop in both total and naive CD4 and CD8 T cells with DPS in aged patients with severe COVID-19. This drop could potentially be explained by increased trafficking to the site of infection or cytokine-storm induced apoptosis as has been reported for other viral diseases (31). To that end, we observe a clear up-regulation of cell death (Fas signaling) pathway in memory CD4 T cells and apoptotic signaling in memory CD8 T cells from aged patients. Thirdly, severe COVID-19 associated metabolites in blood (such as lactic acid) can suppress T cell proliferation (31). Indeed, our data highlights that while relative abundance of Ki67+ proliferating T cell clusters are enriched with severe disease, their overall frequencies are reduced with age. These observations are in line with progressive decrease in plasma levels of T cell proliferation factors IL-2 and IL-7 in aged patients.

In addition to loss of T cells, and suppressed proliferation, there is mounting evidence pointing at T cell dysfunction/exhaustion with severe COVID-19 (34, 48, 54, 55). Circulating T cells in patients with COVID-19 have been described to be functionally exhausted, expressing elevated levels of PD-1 (34, 54). Here, we report gene expression differences in memory CD4 and CD8 T cells indicative of increased T cell activation signatures with severe COVID-19 in the aged subjects. Given previous reports of elevated HLA-DR+ T cells with severe disease (50, 54), we conclude that these observations are suggestive of broad T cell activation, which are exacerbated with age. Furthermore, this hyperactivation of T cells could also explain the depletion of T cells, which is more significant in aged subjects. Functional exhaustion of T cells with severe COVID-19 is further evident from lack of IFN***γ*** production in response to SARS-CoV-2 peptide pools as well as polyclonal stimulation. These findings are consistent with reduced function of CD4 T cells in acute phase of infection following PMA/ionomycin stimulation (56), but are inconsistent with reports of comparable IFN***γ*** CD8 responses or even IFN***γ***, TNFα, and IL-2 CD4 responses following CD3/CD28 stimulation (48, 54). These discrepancies are likely due to differences in the age of the patient, disease severity and the length of stimulation.

Gene expression changes reported here also suggest that CD8 T cells from patients with severe disease exhibit increased cytotoxicity (based on increased expression of *GZMB, PRF1, XCL1, XCL2*) and NK cell markers (*KLRD1*), as previously reported using flow cytometry (25). Importantly, cytotoxicity module scores within CD8 clusters identified by scRNA Seq are significantly higher in aged subjects. However, within the aged cohort with severe disease, patients who succumbed to the disease had attenuated frequencies of *GZMK*^high^ CD8 T cells relative to patients who survived. This observation is line with reports of reduced cytotoxic potential in EM/TEMRA CD8 T cells from severe patients older than 80 yrs of age (36).

Enhanced cytotoxicity with severe disease is also evident by increased frequencies of *GRZMB*+ CD56+ NK cells with severe disease. We also report increased frequencies of KLRG1+ NK cells in aged subjects with severe COVID-19 (57) relative to age-matched HD and young subjects with severe COVID-19. Enhanced activation and functional exhaustion of NK cells have recently been described (30, 36, 57) in the context of severe COVID-19. Our data suggests that this dysregulated state is further exacerbated in aged subjects, with lower frequencies of GranzymeB+ NK cells and reduced expression of *IFNG* and *PRF* gene in the more cytotoxic CD56++ NK cells. In line with these observations, we observed significantly lower levels of circulating Granzyme B in aged patients with severe disease.

Our data show an increase in non-classical monocytes in both young and aged patients with severe disease; however, an increased prevalence of classical monocytes is only evident in young subjects. Consistent with previous observations (29), we report attenuated induction of cytokine/chemokine transcripts in monocytes with severe disease. Surface expression of CCR5, a target for limiting cytokine storms in COVID-19 (58) is significantly elevated early in disease, but wanes with disease progression. We also report an up-regulation of surface CD64 expression, which has also been linked to cytokine release (50, 58), early that wanes with disease progression. Expression of calprotectin (S100A8, S100A9 complex) has been proposed as a factor discriminating mild vs. severe disease (59). Both *S100A8* and *S100A9* were further elevated in monocytes from aged patients with severe disease. Taken together, our studies suggest that while circulating monocytes are not the primary source of cytokine storm in blood, severe COVID-19 is associated with a heightened state of inflammatory signaling in monocytes that is exacerbated in aged subjects. In contrast to monocytes, a significant drop in both myeloid and plasmacytoid dendritic cells in blood is observed with severe disease regardless of age (29, 50). This reduction could be due to recruitment to the lung. Significant induction of factors that regulate hematopoiesis (Flt-3L, G-CSF, IL-33) was only observed in young patients suggesting robust mobilization of myeloid cells in the young group.

As recently described (27, 29, 60), we report a drop in MHC class II molecules (protein and RNA) on both monocytes and DC subsets independent of age. Moreover, HLA-DR expression recovers in young subjects, but remains significantly dampened on monocytes from aged patients. This downregulation of HLA-DR in monocytes coupled with significantly lower CD86 expression suggests skewing of cells into a state of immune tolerance (27). Indeed, our analysis within classical monocytes suggests a preferential trajectory of classical monocytes towards a cluster expressing high levels of *CD163* in aged patients with severe disease. Furthermore, monocytes from patients with severe COVID-19 exhibit attenuated response to TLR stimulation. Similarly, DCs from patients with severe disease also responded poorly to bacterial TLR agonists. Functional impairment of DCs with acute SARS-CoV-2 infection has been recently described (56). These findings suggest that severe disease could potentially lead to innate immune paralysis leaving the host vulnerable to secondary bacterial infections as recently described (61). In contrast to the dampened response following stimulation with bacterial antigens, we report elevated RIG-I signaling module with severe disease, particularly in myeloid DCs in aged subjects. In line with the higher IFN module score, stimulation of both monocytes and DCs with viral agonists targeting TLRs 7/8/9 resulted in higher interferon signal over the course of infection, suggesting innate immune signaling preferentially geared towards antiviral signaling.

Recent studies have highlighted both impairment of type I interferon activity and inflammatory responses with severe COVID-19 (26). Our data suggest systemic elevation of both type I and II interferon in the periphery for all groups. However, cellular expression of interferon signatures varied by disease severity, immune cell type, age, and DPS. Specifically, we observed consistent upregulation of interferon signaling in monocytes, B cells, and NK cells with mild disease and to a lesser degree with severe disease. Within CD8 T cells, while both mild and severe disease triggers an enhanced interferon response, severe disease activates a more robust cytotoxic program. Temporal analysis of interferon responses in NK cells and CD8 T cell clusters in severe disease revealed a late induction of interferon signatures (18-20 DPS), coinciding with progressive decline in plasma IL-10 levels in surviving patients. Mild disease, however, was characterized by up-regulation of feedback regulators such as *IFI44L* (in B cells, DCs, monocytes, NK cells) (62), suggesting mechanisms of efficient control of host innate responses in addition to control of viral replication with mild COVID-19. In contrast, within CD4 T cells and DCs, we observe stronger ISG signaling with severe disease, particularly in the aged subjects, potentially due to delayed viral clearance. Exaggerated innate immunity in the lung, impaired antigen presentation, and lymphopenia are hallmarks of other respiratory viral infections such as influenza and RSV in the aged (17). Our analysis extends these observations and confirms the impairment of innate and adaptive immune responses in blood following severe COVID-19 in aged patients.

We acknowledge several limitations in our study design and implementation. Firstly, we analyzed immune parameters by days post symptom onset, which being self-reported can be rather inaccurate and arbitrary. Secondly, we broadly defined patients with mild disease as ones with a positive PCR test with either no symptoms associated with COVID-19 or a mild disease not requiring extensive (>3 days) hospital stay. Lack of longitudinal samples from patients from this category prevented us from modeling disease dynamics with varied severity. Thirdly, given the nature of this pandemic, there are some biases in patient and healthy donor cohorts. For example, healthy donor subjects were predominantly female (68% in young; 58% in aged) whereas a significant number of patients with severe disease were Hispanic (69% in young; 75% in aged). Additionally, patients in severe categories presented with a wide array of underlying conditions that might have played a role in disease severity/outcome. A significant number of patients were treated with remdesivir, however, there is limited evidence suggesting its role in either immune activation or suppression in blood. Due to limited statistical power, we pooled patients with severe disease at any DPS timepoint into one category for initial analysis before regressing the data with time. Future studies will stratify young and aged patients by clinical scores to identify innate immune correlates of disease severity and identify determinants of disease resolution and survival in patients with severe COVID-19. More importantly, addressing the impact of age on qualitative differences in humoral responses and long-term durability of T cell responses to SARS-CoV-2 would be critical in designing vaccine and therapeutic strategies in the aged population.

## Data Availability

The datasets supporting the conclusions of this article are available on NCBIs Sequence Read Archive (SRA number pending).

## FIGURE LEGENDS

**Supplementary Figure 1: Longitudinal profiling of soluble plasma mediators in mild and severe COVID-19 patients stratified by age**.

(A) Bubble plot comparing plasma concentrations of cytokines, chemokines, and growth factors in healthy donors, young and aged patients with mild and severe disease. The size of the dot represents the log_10_(median concentration) and the color represents the corrected p-value calculated between each infection group and the healthy donors. The # sign denotes factors that are significantly different between the Severe Aged and Severe Young groups. (B) Dot plots representing concentrations (pg/mL) of select cytokines, chemokines and growth factors measured by Luminex assay. *=significance relative to the healthy donors, += significance relative to mild patients, #=significance relative to Young Severe group; *<0.05, **<0.01, ***<0.001, ****<0.0001. (C) MDS plot of proximity scores from a random forest of circulating immune mediators at 1-5 DPS colored by disease severity. (D) Bubble plot of the top five most important predictor variables (1-5 DPS) as ordered by mean decrease in accuracy. The size of the bubble represents the fold-change in concentration vs. healthy controls and the color represents the corrected p-value calculated between each infection group and the healthy donors. (E) MDS plot of proximity scores from a random forest of circulating immune mediators at >10 DPS colored by disease severity. (F) Bubble plot of the top five most important predictor variables (>10 DPS) as ordered by mean decrease in accuracy. The size of the bubble represents the fold-change in concentration vs. healthy controls and the color represents the corrected p-value calculated between each infection group and the healthy donors.

**Supplementary Figure 2: Single cell RNA profiles of PBMC of mild and severe COVID-19 patients stratified by age**.

(A) Longitudinal sampling scheme for scRNA-seq from patients with severe COVID-19. (B) Bar graph comparing the number of cells (log scale) captured from each patient at each timepoint by scRNA-Seq. (C) Bubble plot of identifying genes highly expressed in each cluster determined using the *FindMarkers* function in Seurat. Level of normalized expression is shown using a color scale ranging from low (blue) to high (red). Size of the bubble is representative of the fraction of cells within each cluster expressing the marker. (D) Box plots comparing cell percentages calculated from clusters identified by dimensional reduction of all scRNA-Seq data.

**Supplementary Figure 3: B cell subset frequencies and antibody responses with mild and severe COVID-19 as a function of time**

(A) Bubble plot of identifying genes highly expressed in each B cell cluster determined using Seurat’s *FindMarkers* function. Normalized transcript levels are shown using a color scale ranging from low (blue) to high (red). Size of the bubble is representative of the fraction of cells within each cluster expressing the marker. (B) Stacked bar graph comparing the distribution of the major B cell subsets across each group reported as percentage total cells. (C) Dot plots representing OD values of NP (top) and RBD (bottom) specific IgG and IgM antibodies measured by ELISA at all DPS for indicated age and severity group. *=significant compared to healthy donors, += significant compared to mild patients, #=significant difference between Aged Mild/Severe and Young Mild/Severe groups where */+/#=p<0.05, **=p<0.01, ***=p<0.001, ****=p<0.0001. (D) Bar graphs comparing optical density (OD) of IgG antibodies during early stages (DPS 1-5) of severe infection in young and aged subjects stratified by their ICU status (I-Intubated) and disease outcome.

**Supplementary Figure 4: Phenotypic changes in T cell subsets with mild and severe COVID-19 stratified by age**.

(A) Bubble plot of genes highly expressed in each T cell cluster determined using the *FindMarkers* function in Seurat. Level of normalized expression is shown using a color scale ranging from low (blue) to high (red). Size of the bubble is representative of the fraction of cells within each cluster expressing the marker. (B) Trajectory analysis of CD4 clusters excluding Tregs determined using Slingshot. (C) Box plots comparing frequencies of CD4 TEMRA cells within total cells from each patient identified from single cell analysis of T and NK cell subset. (D) Bar graph showing over-representative GO terms in genes in CD4 TEMRA cells uniquely down-regulated with severe COVID-19 in aged subjects. Gene numbers associated with each term are highlighted next to each term. (E) Violin plot comparing Fas Signaling module scores within memory CD4 T cell clusters. (F) Trajectory analysis of CD8 clusters determined using Slingshot. (G) Violin plots comparing cytotoxicity, cytokine, and interferon module scores among the 4 memory CD8 T cell clusters. (H-K) Box plots comparing frequencies of (H) naive CD8 T cells, (I) IFN CD8 T cells, and (J,K) cytotoxic CD8 T cells within total cells in each patient identified from single cell analysis.

**Supplementary Figure 5: Impact of age on T cell proliferation associated with severe COVID-19**

(A) Violin plots comparing interferon signaling module scores within all memory CD8 T cell clusters. (B) Box plots comparing longitudinal changes in interferon modules in memory CD8 T cells from young and aged subjects with severe COVID-19. X axis represents days post symptom onset (DPS). (C) Box plots comparing frequencies of proliferating T cells within total cells in each patient identified from single cell analysis of T and NK cell subset. (D) Box plots comparing longitudinal changes in IL-2 signaling module in young and aged subjects with severe COVID-19. *=significant compared to healthy donors, += significant compared to mild patients, #=significant difference between Aged Mild/Severe and Young Mild/Severe groups where */+/#=p<0.05, **=p<0.01, ***=p<0.001, ****=p<0.0001.

**Supplementary Figure 6: Enhanced IFN signaling in NK cells with mild and severe COVID-19**

(A) Bubble plot of genes highly expressed in each NK cell cluster determined using the *FindMarkers* function in Seurat. Level of normalized expression is shown using a color scale ranging from low (blue) to high (red). Size of the bubble is representative of the fraction of cells within each cluster expressing the marker. (B) Box plots comparing frequencies of CD8/NK T cells (expressing *CD8A* and *PLCG2*) within total cells in each patient identified from single cell analysis. (C) Linear regression analysis of the percentage CD8/NK T cells calculated from total cells from scRNA-Seq across days post symptom onset (DPS). (D) Frequencies of CD16^high^ NK cells determined by flow cytometry and scRNA-Seq. (E) Violin plots comparing cytokine/chemokine receptor signaling module with the CD16^high^ NK cell cluster. (F) Violin plots comparing interferon signaling module scores in CD56^high^ and the CD16^high^ NK cell subsets. (G) Box plots comparing longitudinal changes in interferon signaling module in CD16^high^ NK cell subset in young and aged subjects with severe COVID-19. X axis represents days post symptom onset (DPS).

**Supplementary Figure 7: Rewiring of myeloid cell phenotypes with severe COVID-19**

(A-B) Dot plots comparing median fluorescence intensities of HLA-DR and co-stimulation marker CD86 in (A) classical and non-classical (NC) monocytes and (B) myeloid DCs (mDC) and plasmacytoid DCs (pDC). (C) UMAP projection of monocytes and DCs reclustered from the main UMAP. Major subsets of monocytes and DCs are highlighted. (D) Bubble plot of genes highly expressed in each cluster determined using Seurat’s *FindMarkers* function. Level of normalized expression is shown using a color scale ranging from low (blue) to high (red). Size of the bubble is representative of the fraction of cells within each cluster expressing the marker. (E) Violin plot comparing changes in MHC class II RNA module scores within classical and non-classical monocytes. (F) Stacked bar graph quantifying proportional changes in myeloid cell subsets with mild and severe COVID-19. (G) Box plots comparing relative frequencies of monocyte clusters that increase/decrease significantly with COVID-19. (H) Bar graphs comparing key monocyte surface markers in healthy donors (HD) and severe COVID-19 patients sampled during different stages of acute infection post symptom onset. Regression panel below each bar graph compares progression of the marker over the course of acute infection. *=significant compared to healthy donors, += significant compared to mild patients, #=significant difference between Aged Mild/Severe and Young Mild/Severe groups where */+/#=p<0.05, **=p<0.01, ***=p<0.001, ****=p<0.0001.

**Supplementary Figure 8: Compromised monocyte response to secondary insult with severe COVID-19**

(A) Functional enrichment of genes differentially expressed in non-classical monocytes following mild and severe COVID-19 Color and size of the bubble represents statistical significance and number of genes respectively. (B) Bubble plot showing gene expression changes in myeloid DCs from young and aged patients with mild and severe COVID-19. Size of the bubble is representative of the fraction of cells within each cluster expressing the marker. (C) Comparison of changes in RIG-I signaling module scores with COVID-19 in DC subsets.

## Data availability

The datasets supporting the conclusions of this article are available on NCBI’s Sequence Read Archive (SRA# pending).

## Competing interests

Alpesh Amin reported serving as PI or co-I of clinical trials sponsored by NIH/NIAID, NeuroRx Pharma, Pulmotect, Blade Therapeutics, Novartis, Takeda, Humanigen, Eli Lilly, PTC Therapeutics, OctaPharma, Fulcrum Therapeutics, Alexion. He has served as speaker or consultant for BMS, Pfizer, BI, Portola, Sunovion, Mylan/Theravance, Salix, Alexion, AstraZeneca, Novartis, Nabriva, Paratek, Bayer, Tetraphase, Achogen LaJolla, Millenium, Ferring, PeraHealth, HeartRite, Aseptiscope, Sprightly.

## Funding

This study was supported by the National Center for Research Resources and the National Center for Advancing Translational Sciences, National Institutes of Health, through Grant <u>UL1 TR001414</u>. S.A.L is supported by NIH F31 <u>A028704</u>. The content is solely the responsibility of the authors and does not necessarily represent the official views of the NIH.

## Author Contributions

S.A.L., S.S., and I.M. conceived and designed the experiments. S.A.L, S.S, M.Z, B.D, M.C, and A.J performed the experiments. S.A.L, S.S, M.Z, B.D, M.C, and N.R analyzed the data. S.A.L., S.S. and I.M. wrote the paper. All authors have read and approved the final draft of the manuscript.

## Acknowledgements

We are grateful to all participants in this study. We thank Dr. Jennifer Atwood for assistance with sorting and imaging flow cytometry in the flow cytometry core at the Institute for Immunology, UCI. We thank Dr. Melanie Oakes from UCI Genomics and High-Throughput Facility for assistance with 10X library preparation and sequencing. Aspects of experimental design figures were generated using graphics from Biorender.com. We wish to acknowledge the support of the Chao Family Comprehensive Cancer Center resources, supported by the National Cancer Institute of the National Institutes of Health under award number P30CA062203.

## REFERENCES

1. Xie J, Tong Z, Guan X, Du B, Qiu H. Clinical Characteristics of Patients Who Died of Coronavirus Disease 2019 in China. JAMA Netw Open. 2020;3(4):e205619.

2. Nishiura H, Kobayashi T, Miyama T, Suzuki A, Jung SM, Hayashi K, et al. Estimation of the asymptomatic ratio of novel coronavirus infections (COVID-19). Int J Infect Dis. 2020;94:154–5.

3. Mizumoto K, Chowell G. Transmission potential of the novel coronavirus (COVID-19) onboard the diamond Princess Cruises Ship, 2020. Infect Dis Model. 2020;5:264–70.

4. Nabors C, Sridhar A, Hooda U, Lobo SA, Levine A, Frishman WH, et al. Characteristics and Outcomes of Patients 80 Years and Older Hospitalized With Coronavirus Disease 2019 (COVID-19). Cardiol Rev. 2021;29(1):39–42.

5. Grasselli G, Zangrillo A, Zanella A, Antonelli M, Cabrini L, Castelli A, et al. Baseline Characteristics and Outcomes of 1591 Patients Infected With SARS-CoV-2 Admitted to ICUs of the Lombardy Region, Italy. JAMA. 2020;323(16):1574–81.

6. Mueller AL, McNamara MS, Sinclair DA. Why does COVID-19 disproportionately affect older people? Aging (Albany NY). 2020;12(10):9959–81.

7. Levin AT, Hanage WP, Owusu-Boaitey N, Cochran KB, Walsh SP, Meyerowitz-Katz G. Assessing the age specificity of infection fatality rates for COVID-19: systematic review, meta-analysis, and public policy implications. Eur J Epidemiol. 2020;35(12):1123–38.

8. Li J, Gong X, Wang Z, Chen R, Li T, Zeng D, et al. Clinical features of familial clustering in patients infected with 2019 novel coronavirus in Wuhan, China. Virus Res. 2020;286:198043.

9. Wang D, Hu B, Hu C, Zhu F, Liu X, Zhang J, et al. Clinical Characteristics of 138 Hospitalized Patients With 2019 Novel Coronavirus-Infected Pneumonia in Wuhan, China. JAMA. 2020;323(11):1061–9.

10. Wu J, Li J, Zhu G, Zhang Y, Bi Z, Yu Y, et al. Clinical Features of Maintenance Hemodialysis Patients with 2019 Novel Coronavirus-Infected Pneumonia in Wuhan, China. Clin J Am Soc Nephrol. 2020;15(8):1139–45.

11. Docherty AB, Harrison EM, Green CA, Hardwick HE, Pius R, Norman L, et al. Features of 20?133 UK patients in hospital with covid-19 using the ISARIC WHO Clinical Characterisation Protocol: prospective observational cohort study. BMJ. 2020;369:m1985.

12. Verity R, Okell LC, Dorigatti I, Winskill P, Whittaker C, Imai N, et al. Estimates of the severity of coronavirus disease 2019: a model-based analysis. Lancet Infect Dis. 2020;20(6):669–77.

13. Liu K, Chen Y, Lin R, Han K. Clinical features of COVID-19 in elderly patients: A comparison with young and middle-aged patients. J Infect. 2020;80(6):e14–e8.

14. Hazeldine J, Lord JM. Immunesenescence: A Predisposing Risk Factor for the Development of COVID-19? Front Immunol. 2020;11:573662.

15. Perrotta F, Corbi G, Mazzeo G, Boccia M, Aronne L, D’Agnano V, et al. COVID-19 and the elderly: insights into pathogenesis and clinical decision-making. Aging Clin Exp Res. 2020;32(8):1599–608.

16. Aiello A, Farzaneh F, Candore G, Caruso C, Davinelli S, Gambino CM, et al. Immunosenescence and Its Hallmarks: How to Oppose Aging Strategically? A Review of Potential Options for Therapeutic Intervention. Front Immunol. 2019;10:2247.

17. Chen J, Kelley WJ, Goldstein DR. Role of Aging and the Immune Response to Respiratory Viral Infections: Potential Implications for COVID-19. J Immunol. 2020;205(2):313– 20.

18. Akbar AN, Gilroy DW. Aging immunity may exacerbate COVID-19. Science. 2020;369(6501):256–7.

19. Blanco-Melo D, Nilsson-Payant BE, Liu WC, Uhl S, Hoagland D, Møller R, et al. Imbalanced Host Response to SARS-CoV-2 Drives Development of COVID-19. Cell. 2020;181(5):1036-45.e9.

20. Laing AG, Lorenc A, Del Molino Del Barrio I, Das A, Fish M, Monin L, et al. A dynamic COVID-19 immune signature includes associations with poor prognosis. Nat Med. 2020;26(10):1623–35.

21. Lucas C, Wong P, Klein J, Castro TBR, Silva J, Sundaram M, et al. Longitudinal analyses reveal immunological misfiring in severe COVID-19. Nature. 2020;584(7821):463–9.

22. Vabret N, Britton GJ, Gruber C, Hegde S, Kim J, Kuksin M, et al. Immunology of COVID-19: Current State of the Science. Immunity. 2020;52(6):910–41.

23. Del Valle DM, Kim-Schulze S, Huang HH, Beckmann ND, Nirenberg S, Wang B, et al. An inflammatory cytokine signature predicts COVID-19 severity and survival. Nat Med. 2020;26(10):1636–43.

24. Giamarellos-Bourboulis EJ, Netea MG, Rovina N, Akinosoglou K, Antoniadou A, Antonakos N, et al. Complex Immune Dysregulation in COVID-19 Patients with Severe Respiratory Failure. Cell Host Microbe. 2020;27(6):992-1000.e3.

25. Kuri-Cervantes L, Pampena MB, Meng W, Rosenfeld AM, Ittner CAG, Weisman AR, et al. Comprehensive mapping of immune perturbations associated with severe COVID-19. Sci Immunol. 2020;5(49).

26. Hadjadj J, Yatim N, Barnabei L, Corneau A, Boussier J, Smith N, et al. Impaired type I interferon activity and inflammatory responses in severe COVID-19 patients. Science. 2020;369(6504):718–24.

27. Schulte-Schrepping J, Reusch N, Paclik D, Baßler K, Schlickeiser S, Zhang B, et al. Severe COVID-19 Is Marked by a Dysregulated Myeloid Cell Compartment. Cell. 2020;182(6):1419-40.e23.

28. Osman M, Faridi RM, Sligl W, Shabani-Rad MT, Dharmani-Khan P, Parker A, et al. Impaired natural killer cell counts and cytolytic activity in patients with severe COVID-19. Blood Adv. 2020;4(20):5035–9.

29. Wilk AJ, Rustagi A, Zhao NQ, Roque J, Martĺnez-Colón GJ, McKechnie JL, et al. A single-cell atlas of the peripheral immune response in patients with severe COVID-19. Nat Med. 2020;26(7):1070–6.

30. Maucourant C, Filipovic I, Ponzetta A, Aleman S, Cornillet M, Hertwig L, et al. Natural killer cell immunotypes related to COVID-19 disease severity. Sci Immunol. 2020;5(50).

31. Tan L, Wang Q, Zhang D, Ding J, Huang Q, Tang YQ, et al. Lymphopenia predicts disease severity of COVID-19: a descriptive and predictive study. Signal Transduct Target Ther. 2020;5(1):33.

32. Chen Z, John Wherry E. T cell responses in patients with COVID-19. Nat Rev Immunol. 2020;20(9):529–36.

33. Zheng M, Gao Y, Wang G, Song G, Liu S, Sun D, et al. Functional exhaustion of antiviral lymphocytes in COVID-19 patients. Cell Mol Immunol. 2020;17(5):533–5.

34. Diao B, Wang C, Tan Y, Chen X, Liu Y, Ning L, et al. Reduction and Functional Exhaustion of T Cells in Patients With Coronavirus Disease 2019 (COVID-19). Front Immunol. 2020;11:827.

35. Mazzoni A, Salvati L, Maggi L, Capone M, Vanni A, Spinicci M, et al. Impaired immune cell cytotoxicity in severe COVID-19 is IL-6 dependent. J Clin Invest. 2020;130(9):4694–703.

36. Westmeier J, Paniskaki K, Karaköse Z, Werner T, Sutter K, Dolff S, et al. Impaired Cytotoxic CD8. mBio. 2020;11(5).

37. Zheng Y, Liu X, Le W, Xie L, Li H, Wen W, et al. A human circulating immune cell landscape in aging and COVID-19. Protein Cell. 2020;11(10):740–70.

38. Butler A, Hoffman P, Smibert P, Papalexi E, Satija R. Integrating single-cell transcriptomic data across different conditions, technologies, and species. Nat Biotechnol. 2018;36(5):411–20.

39. Zheng GX, Terry JM, Belgrader P, Ryvkin P, Bent ZW, Wilson R, et al. Massively parallel digital transcriptional profiling of single cells. Nat Commun. 2017;8:14049.

40. Street K, Risso D, Fletcher RB, Das D, Ngai J, Yosef N, et al. Slingshot: cell lineage and pseudotime inference for single-cell transcriptomics. BMC Genomics. 2018;19(1):477.

41. Singer M, Wang C, Cong L, Marjanovic ND, Kowalczyk MS, Zhang H, et al. A Distinct Gene Module for Dysfunction Uncoupled from Activation in Tumor-Infiltrating T Cells. Cell. 2017;171(5):1221–3.

42. Zhou Y, Zhou B, Pache L, Chang M, Khodabakhshi AH, Tanaseichuk O, et al. Metascape provides a biologist-oriented resource for the analysis of systems-level datasets. Nat Commun. 2019;10(1):1523.

43. Gold JAW, Rossen LM, Ahmad FB, Sutton P, Li Z, Salvatore PP, et al. Race, Ethnicity, and Age Trends in Persons Who Died from COVID-19 - United States, May-August 2020. MMWR Morb Mortal Wkly Rep. 2020;69(42):1517–21.

44. Lang FM, Lee KM, Teijaro JR, Becher B, Hamilton JA. GM-CSF-based treatments in COVID-19: reconciling opposing therapeutic approaches. Nat Rev Immunol. 2020;20(8):507–14.

45. Culley FJ, Pennycook AM, Tregoning JS, Dodd JS, Walzl G, Wells TN, et al. Role of CCL5 (RANTES) in viral lung disease. J Virol. 2006;80(16):8151–7.

46. Chen Y, Wang J, Liu C, Su L, Zhang D, Fan J, et al. IP-10 and MCP-1 as biomarkers associated with disease severity of COVID-19. Mol Med. 2020;26(1):97.

47. Zhao Y, Qin L, Zhang P, Li K, Liang L, Sun J, et al. Longitudinal COVID-19 profiling associates IL-1RA and IL-10 with disease severity and RANTES with mild disease. JCI Insight. 2020;5(13).

48. De Biasi S, Meschiari M, Gibellini L, Bellinazzi C, Borella R, Fidanza L, et al. Marked T cell activation, senescence, exhaustion and skewing towards TH17 in patients with COVID-19 pneumonia. Nat Commun. 2020;11(1):3434.

49. Pacha O, Sallman MA, Evans SE. COVID-19: a case for inhibiting IL-17? Nat Rev Immunol. 2020;20(6):345–6.

50. Mann ER, Menon M, Knight SB, Konkel JE, Jagger C, Shaw TN, et al. Longitudinal immune profiling reveals key myeloid signatures associated with COVID-19. Sci Immunol. 2020;5(51).

51. Bernardes JP, Mishra N, Tran F, Bahmer T, Best L, Blase JI, et al. Longitudinal Multi-omics Analyses Identify Responses of Megakaryocytes, Erythroid Cells, and Plasmablasts as Hallmarks of Severe COVID-19. Immunity. 2020;53(6):1296-314.e9.

52. Yu K, He J, Wu Y, Xie B, Liu X, Wei B, et al. Dysregulated adaptive immune response contributes to severe COVID-19. Cell Res. 2020;30(9):814–6.

53. Agrati C, Sacchi A, Bordoni V, Cimini E, Notari S, Grassi G, et al. Expansion of myeloid-derived suppressor cells in patients with severe coronavirus disease (COVID-19). Cell Death Differ. 2020;27(11):3196–207.

54. Mathew D, Giles JR, Baxter AE, Oldridge DA, Greenplate AR, Wu JE, et al. Deep immune profiling of COVID-19 patients reveals distinct immunotypes with therapeutic implications. Science. 2020;369(6508).

55. Kalfaoglu B, Almeida-Santos J, Tye CA, Satou Y, Ono M. T-Cell Hyperactivation and Paralysis in Severe COVID-19 Infection Revealed by Single-Cell Analysis. Front Immunol. 2020;11:589380.

56. Zhou R, To KK, Wong YC, Liu L, Zhou B, Li X, et al. Acute SARS-CoV-2 Infection Impairs Dendritic Cell and T Cell Responses. Immunity. 2020;53(4):864-77.e5.

57. Varchetta S, Mele D, Oliviero B, Mantovani S, Ludovisi S, Cerino A, et al. Unique immunological profile in patients with COVID-19. Cell Mol Immunol. 2020.

58. Patterson BK, Seethamraju H, Dhody K, Corley MJ, Kazempour K, Lalezari J, et al. CCR5 inhibition in critical COVID-19 patients decreases inflammatory cytokines, increases CD8 T-cells, and decreases SARS-CoV2 RNA in plasma by day 14. Int J Infect Dis. 2020;103:25–32.

59. Silvin A, Chapuis N, Dunsmore G, Goubet AG, Dubuisson A, Derosa L, et al. Elevated Calprotectin and Abnormal Myeloid Cell Subsets Discriminate Severe from Mild COVID-19. Cell. 2020;182(6):1401-18.e18.

60. Arunachalam PS, Wimmers F, Mok CKP, Perera RAPM, Scott M, Hagan T, et al. Systems biological assessment of immunity to mild versus severe COVID-19 infection in humans. Science. 2020;369(6508):1210–20.

61. Ripa M, Galli L, Poli A, Oltolini C, Spagnuolo V, Mastrangelo A, et al. Secondary infections in patients hospitalized with COVID-19: incidence and predictive factors. Clin Microbiol Infect. 2020.

62. DeDiego ML, Martinez-Sobrido L, Topham DJ. Novel Functions of IFI44L as a Feedback Regulator of Host Antiviral Responses. J Virol. 2019;93(21).

