## Supplemental Materials for "Longitudinal analyses reveal age-specific immune correlates of COVID-19 severity"

Supplemental Figure 1

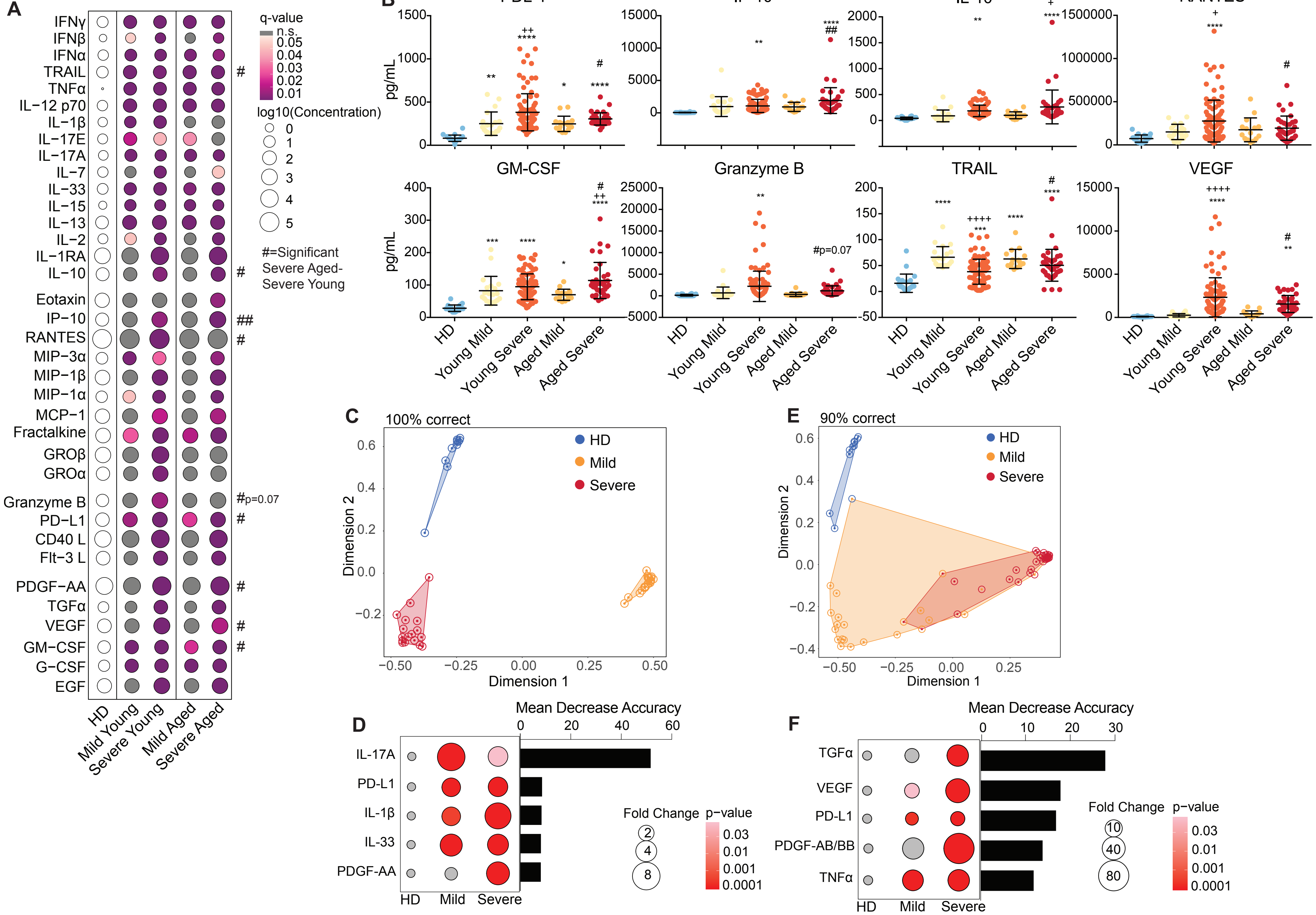

Supplemental Figure 2

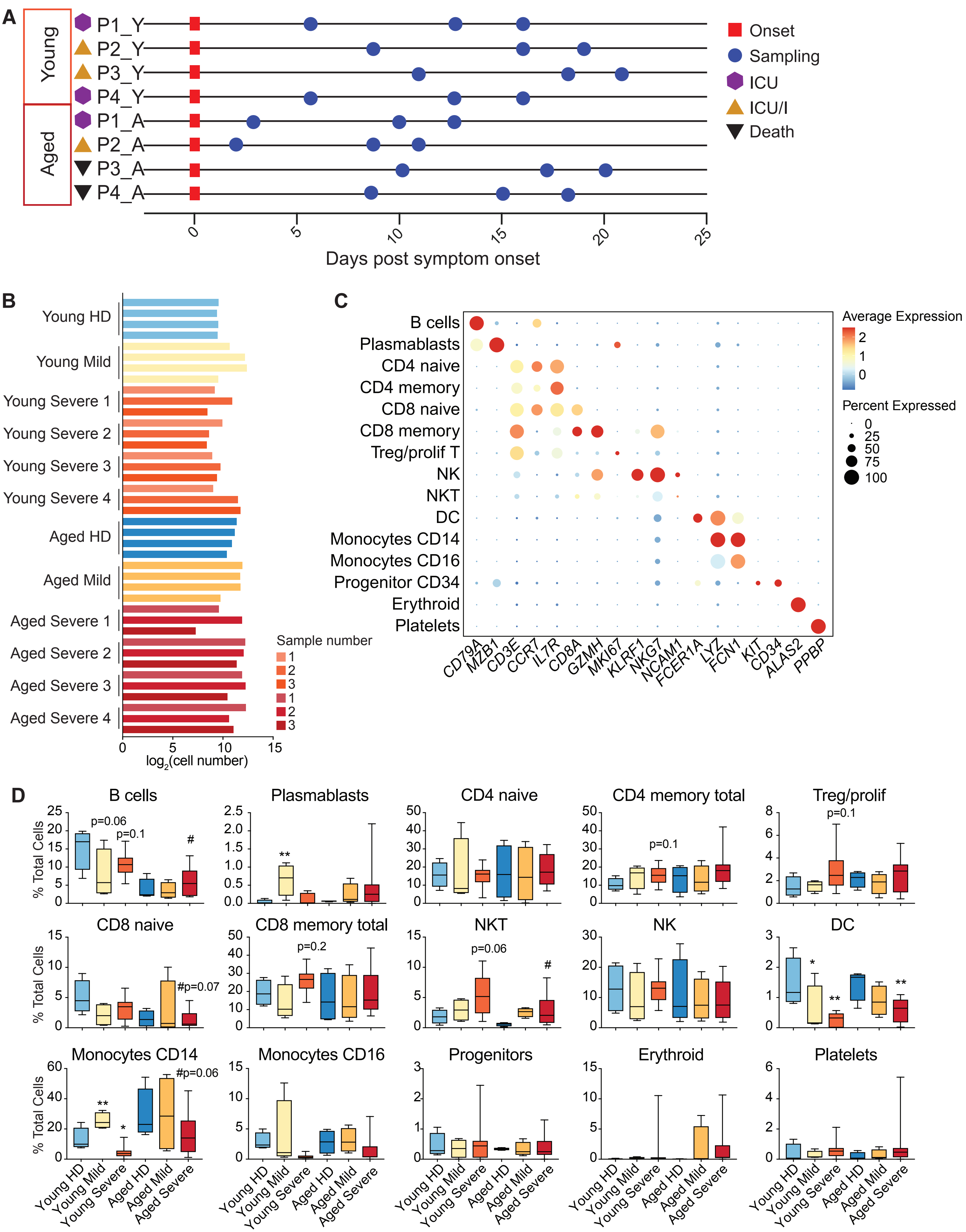

Supplemental Figure 3

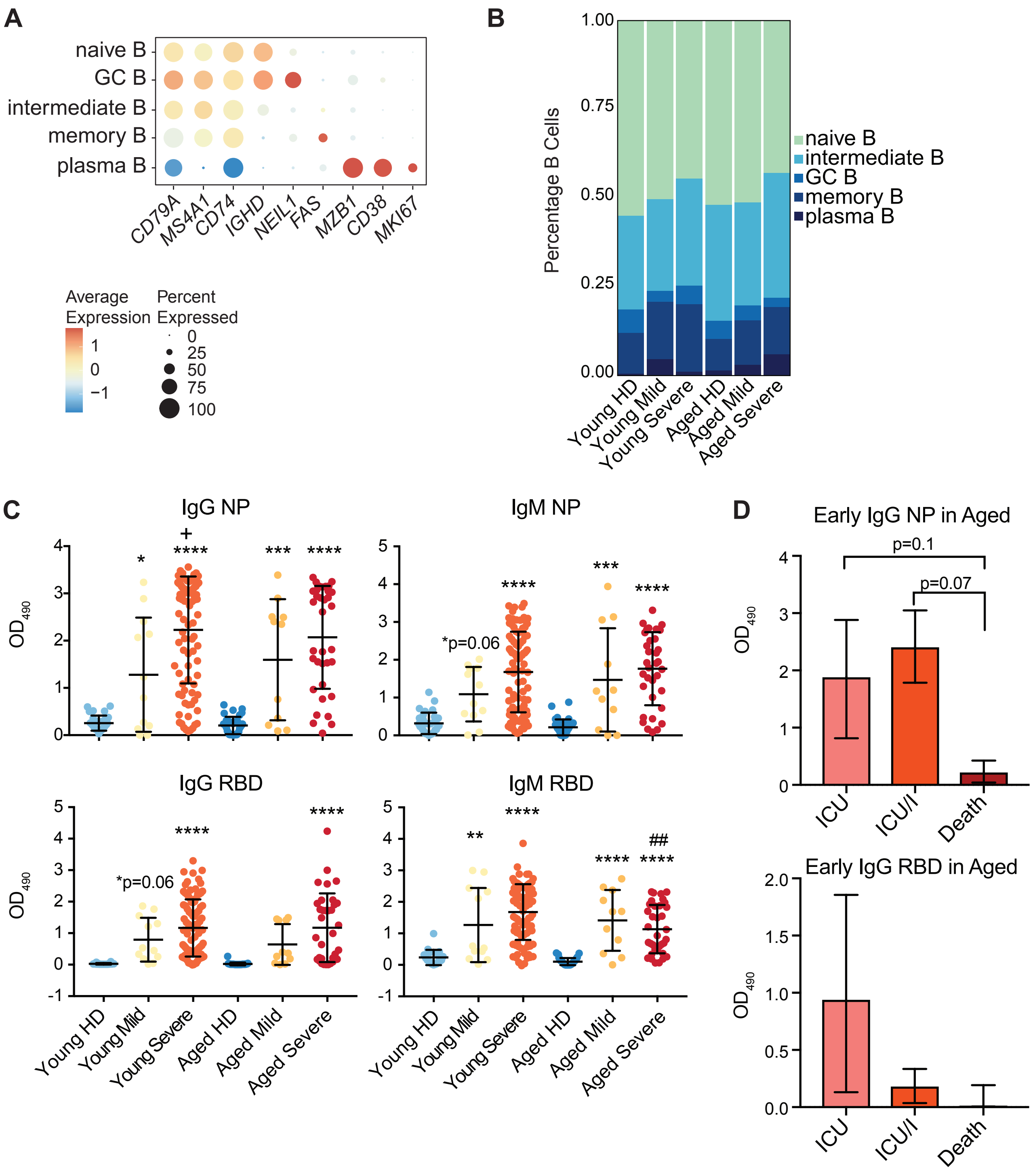

Supplemental Figure 4

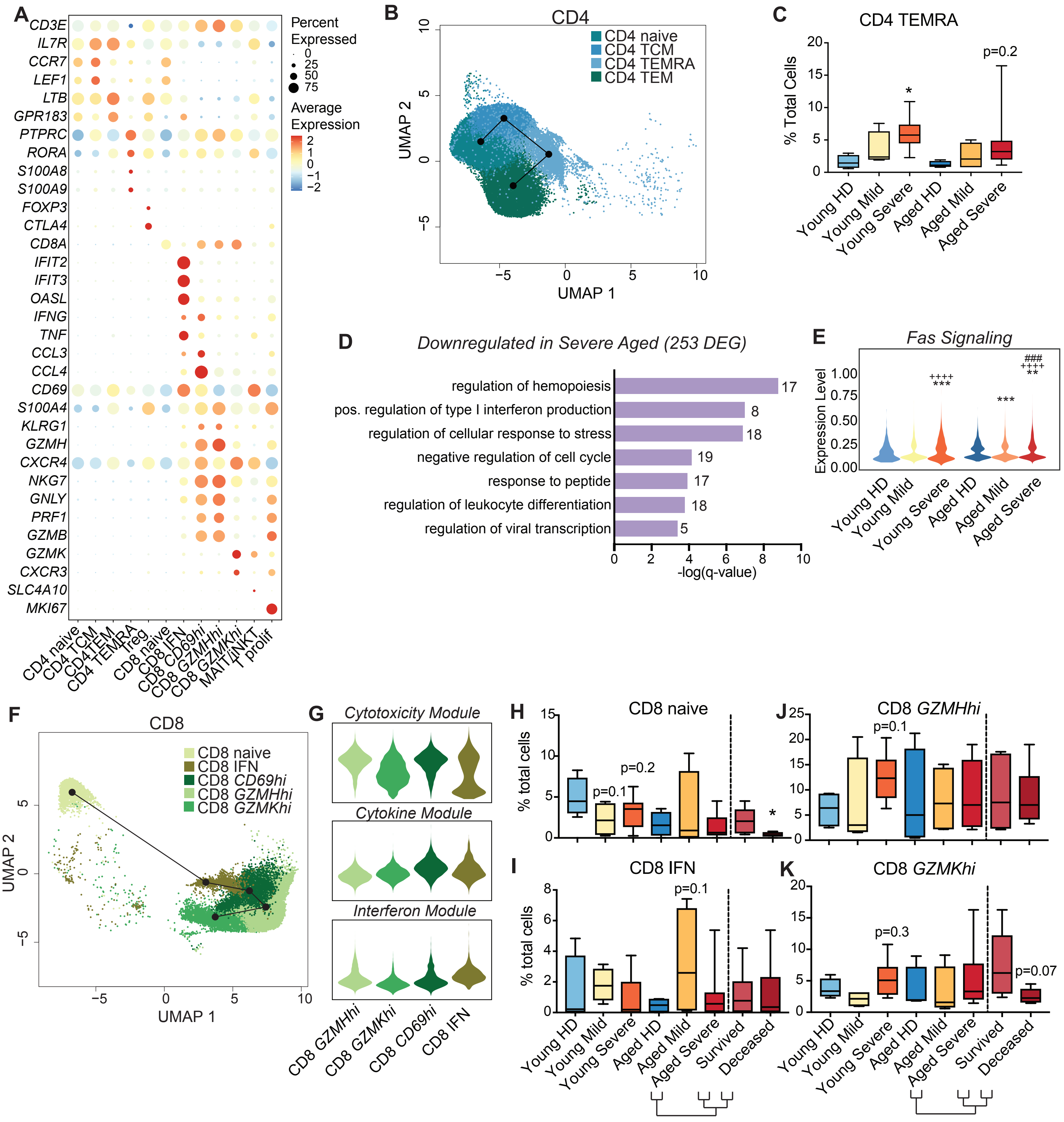

Supplemental Figure 5

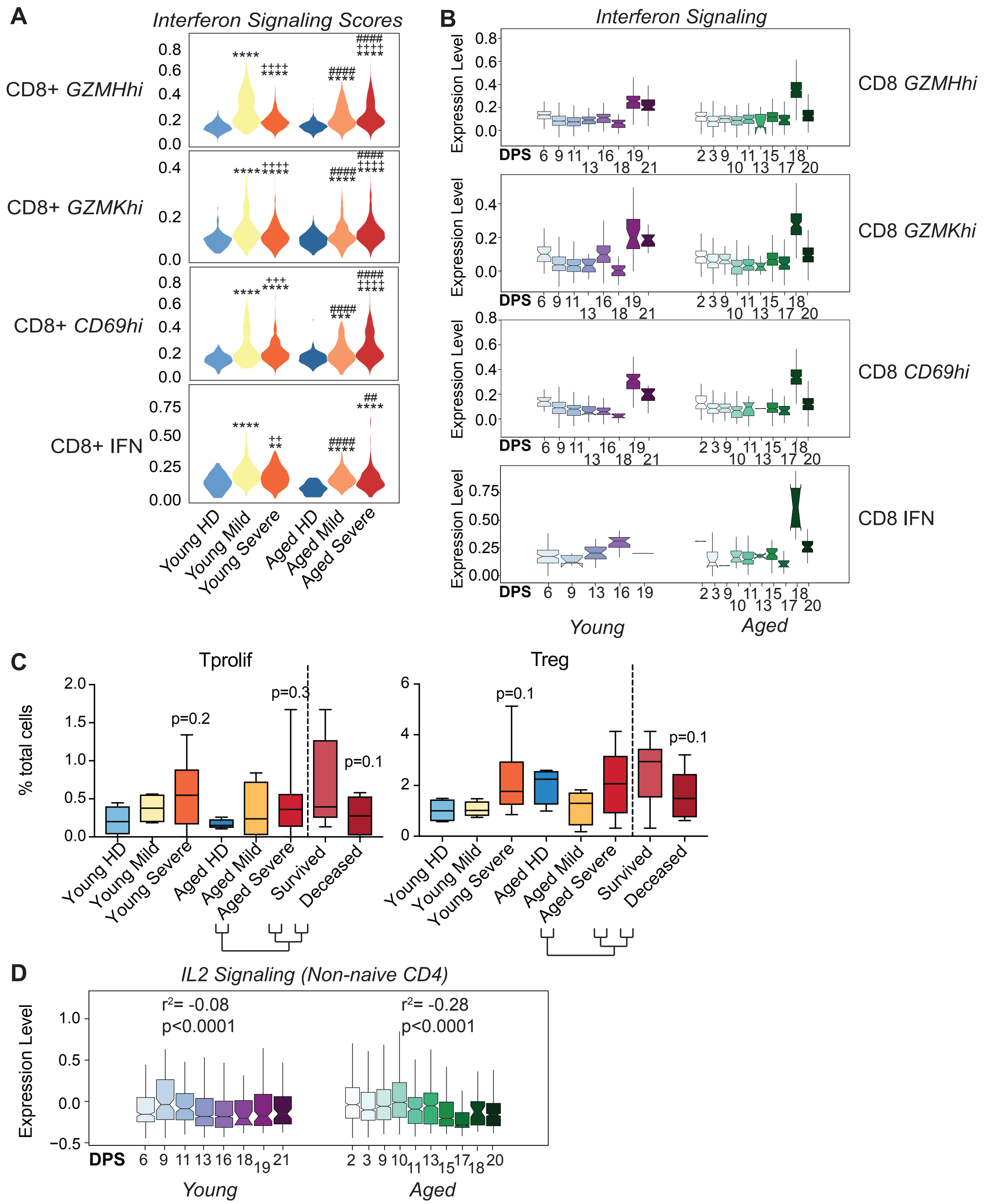

Supplemental Figure 6

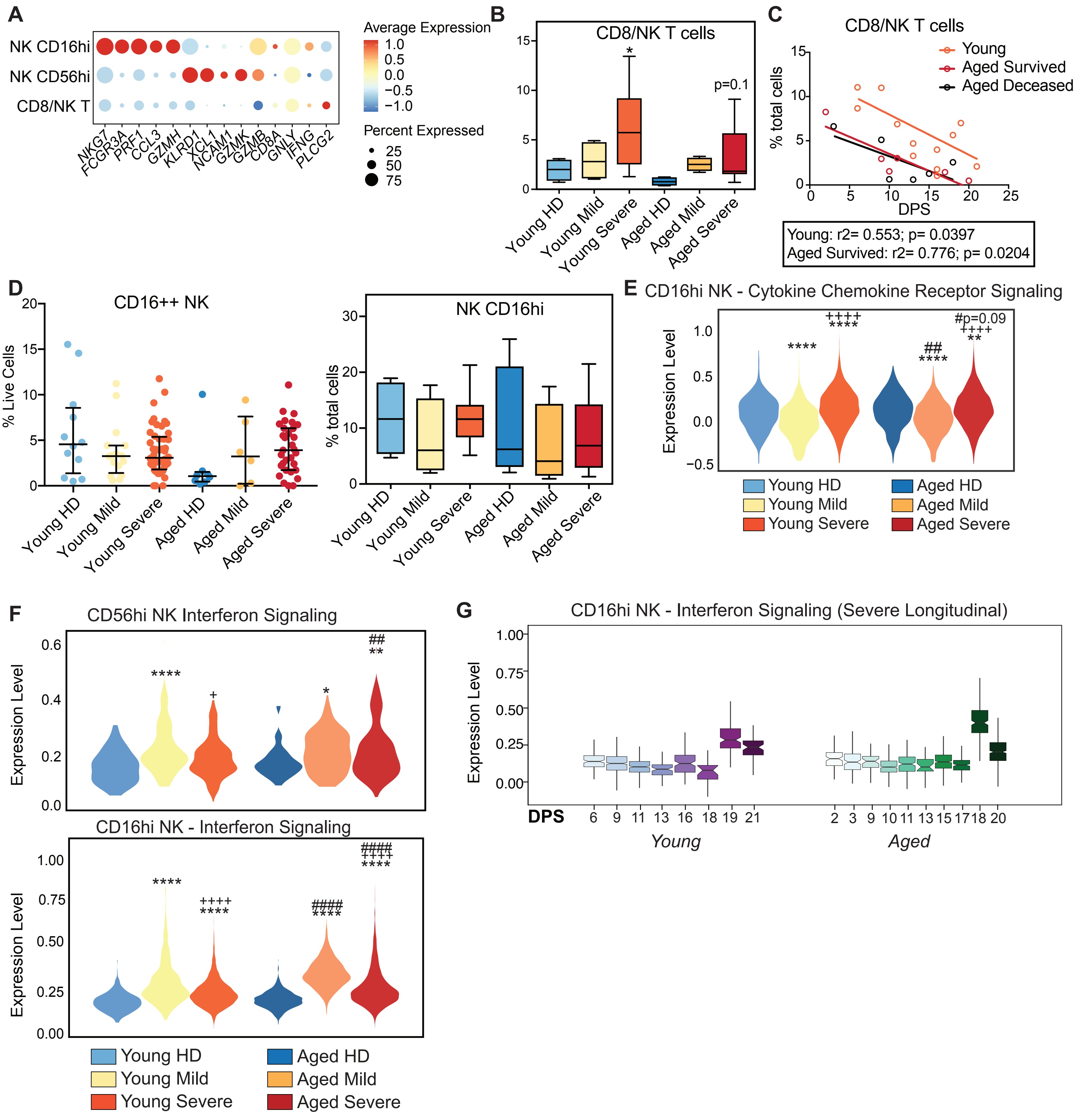

Supplemental Figure 7

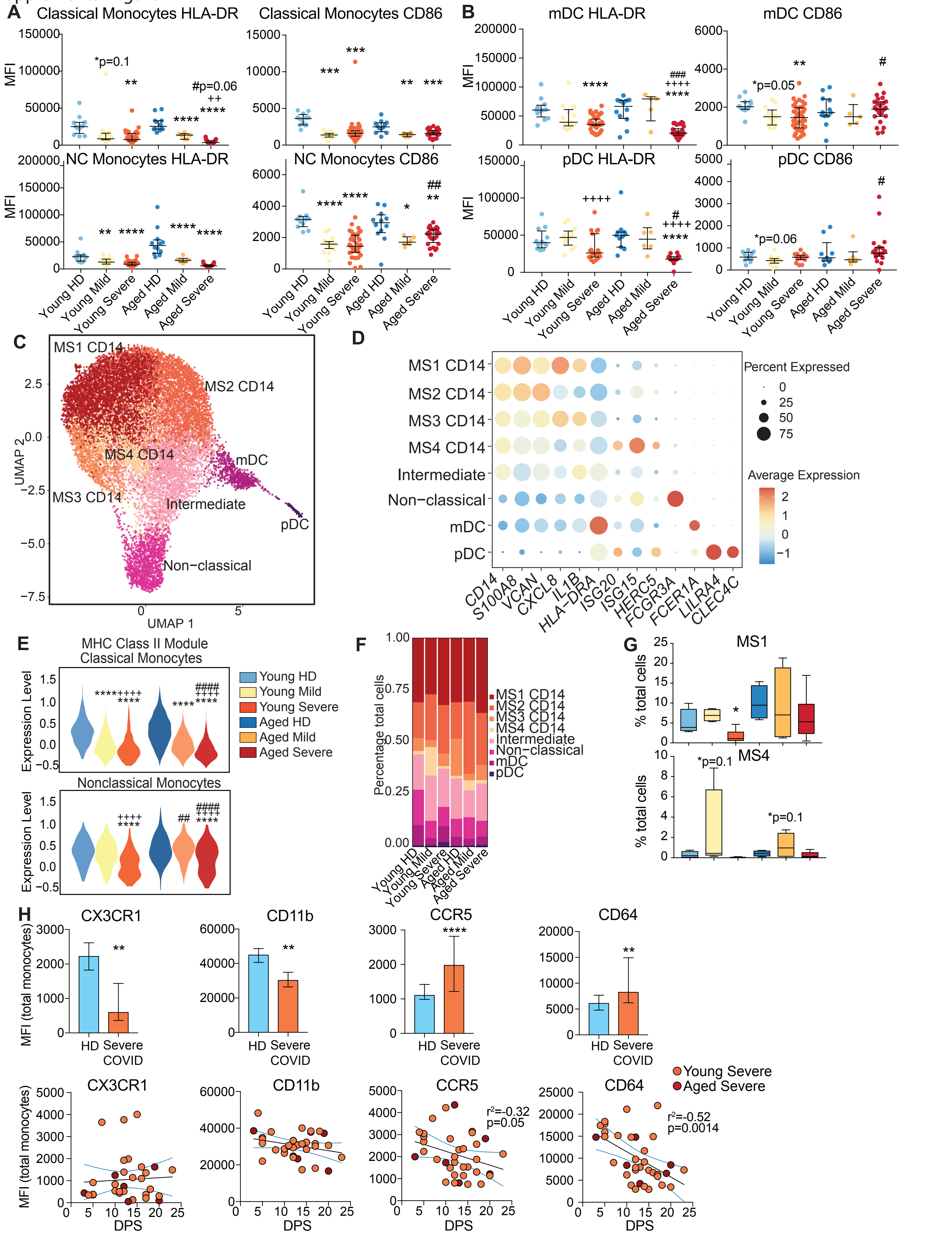

Supplemental Figure 8

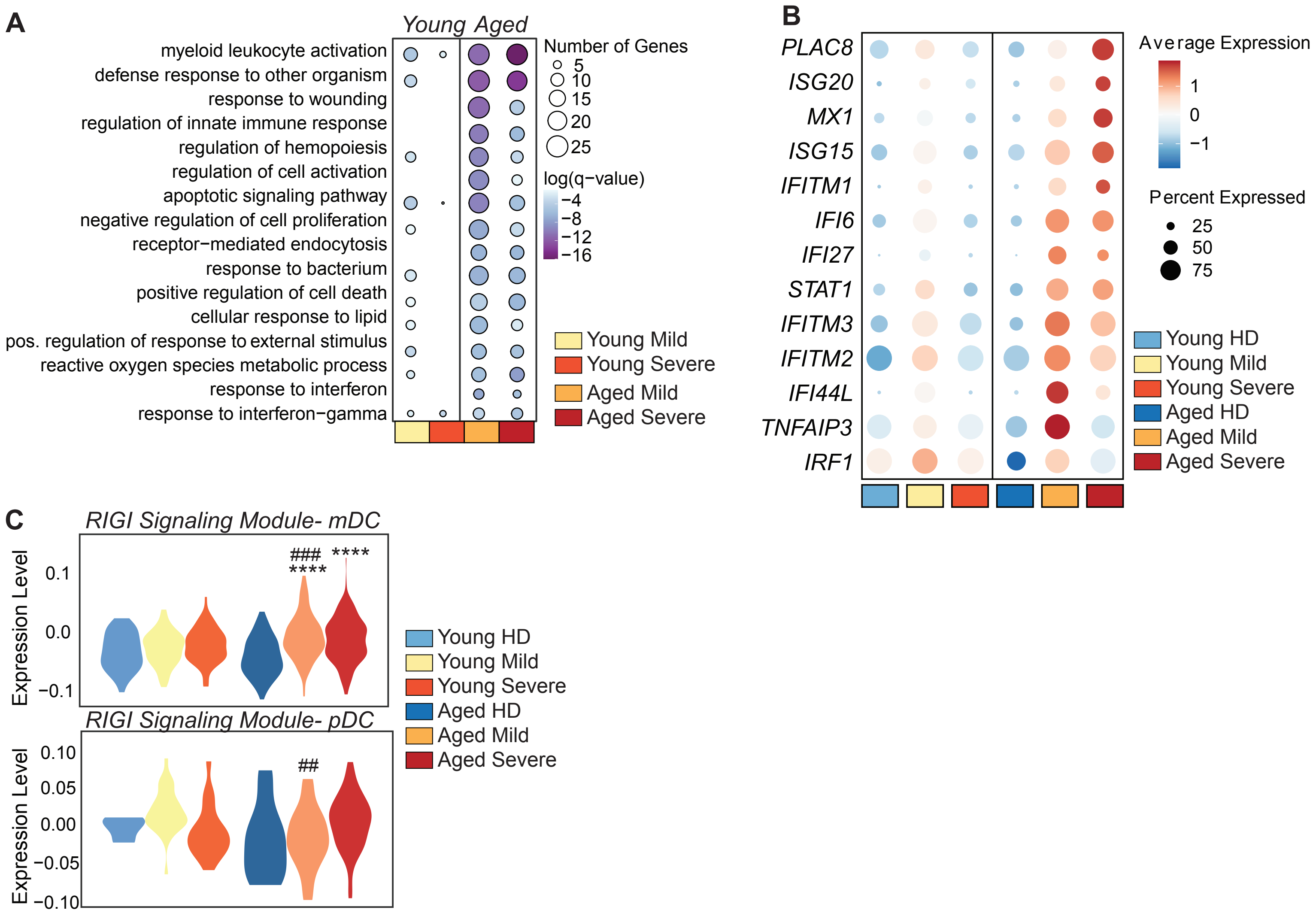

Supplementary Table 1

| | | Patient | Severity | Race | Age | Gender | Comorbidities |
|---|---|---|---|---|---|---|---|
| Healthy Donors | Young | HDY1 | HC | Other | 20's | Female | |
| | | HDY2 | HC | Hispanic | 20's | Female | |
| | | HDY3 | HC | Other | 30's | Male | |
| | | HDY4 | HC | Asian | 30's | Female | |
| | | HDY5 | HC | Caucasian | 30's | Female | |
| | | HDY6 | HC | Other | 30's | Male | |
| | | HDY7 | HC | Other | 30's | Male | |
| | | HDY8 | HC | Other | 30's | Female | |
| | | HDY9 | HC | Other | 30's | Female | |
| | | HDY10 | HC | Hispanic | 30's | Male | |
| | | HDY11 | HC | Caucasian | 30's | Female | |
| | | HDY12 | HC | Other | 30's | Female | |
| | | HDY13 | HC | Other | 30's | Male | |
| | | HDY14 | HC | Caucasian | 40's | Female | |
| | | HDY15 | HC | Caucasian | 40's | Male | |
| | | HDY16 | HC | Caucasian | 40's | Female | |
| | | HDY17 | HC | Asian | 40's | Female | |
| | | HDY18 | HC | Hispanic | 40's | Male | |
| | | HDY19 | HC | Hispanic | 50's | Female | |
| | | HDY20 | HC | Caucasian | 50's | Female | |
| | | HDY21 | HC | Hispanic | 50's | Female | |
| | | HDY22 | HC | Caucasian | 50's | Female | HTN |
| | | HDY23 | HC | Caucasian | 50's | Female | |
| | | HDY24 | HC | Caucasian | 20's | Female | |
| | | HDY25 | HC | Caucasian | 30's | Male | |
| | | HDY26 | HC | Caucasian | 30's | Female | |
| | | HDY27 | HC | Caucasian | 30's | Female | |
| | | HDY28 | HC | Other | 30's | Female | |
| | | HDY29 | HC | Caucasian | 30's | Female | |
| | | HDY30 | HC | Caucasian | 30's | Female | |
| | | HDY31 | HC | Caucasian | 30's | Female | |
| | | HDY32 | HC | Caucasian | 40's | Female | |
| | | HDY33 | HC | Caucasian | 50's | Female | |
| | | HDY34 | HC | Hispanic | 30's | Male | |
| | | HDY35 | HC | Other | 40's | Male | |
| | | HDY36 | HC | White | 20's | Male | |
| | | HDY37 | HC | White | 30's | Male | |
| | | HDA1 | HC | Caucasian | 60's | Female | |
| | | HDA2 | HC | Caucasian | 60's | Female | |
| | | HDA3 | HC | Caucasian | 60's | Female | |

| | | | | | | | |
|---|---|---|---|---|---|---|---|
| | | HDA4 | HC | caucasian | 60's | Male | |
| | | HDA5 | HC | caucasian | 60's | Female | |
| | Aged | HDA6 | HC | caucasian | 60's | Female | |
| | | HDA7 | HC | caucasian | 70's | Male | |
| | | HDA8 | HC | caucasian | 70's | Female | |
| | | HDA9 | HC | caucasian | 70's | Male | |
| | | HDA10 | HC | caucasian | 80's | Female | |
| | | HDA11 | HC | caucasian | 80's | Male | |
| | | HDA12 | HC | caucasian | 80's | Male | |
| | | MY1 | Asymptomatic, Ward | White | 20's | Female | HTN |
| | | MY2 | Asymptomatic, Ward | Hispanic | 40's | Male | |
| | | MY3 | Mild | Unknown | 40's | Female | |
| | | MY4 | Asymptomatic, Ward | Hispanic | 40's | Female | Diabetes |
| | Young | MY5 | Asymptomatic, Ward | White | 40's | Female | iabetes, HTN, CVD, CKD/ESR |
| | | MY6 | Asymptomatic, Ward | Hispanic | 50's | Male | Diabetes, HTN, Obesity |
| | | MY7 | Mild, Ward | White | 50's | Male | HTN, Obesity |
| | | MY8 | Asymptomatic, Ward | Pacific Islander | 50's | Male | iabetes, HTN, A/COPD, Obesi |
| | | MY9 | Mild, Ward | Hispanic | 50's | Male | Obesity |
| Mild/Asymptomatic | | MY10 | Mild, Ward | Hispanic | 50's | Male | Diabetes, HTN, Obesity |
| | | MA1 | Mild, Ward | Hispanic | 60's | Male | Diabetes, HTN, CKD/ESRD |
| | | MA2 | Mild, Ward | Hispanic | 60's | Female | Diabetes, HTN |
| | | MA3 | Asymptomatic, Ward | Hispanic | 60's | Male | Diabetes, HTN |
| | | MA4 | Mild, ER visit only | Hispanic | 60's | Male | Diabetes, HTN, Obesity |
| | Aged | MA5 | Mild, Ward | White | 60's | Male | Diabetes, HTN, Obesity |
| | | MA6 | Asymptomatic, Death | Asian | 70's | Male | HTN, CKD/ESRD |
| | | MA7 | Mild, Ward | Hispanic | 70's | Male | HTN,Obesity |
| | | MA8 | Asymptomatic, Ward | White | 70's | Female | Obesity |
| | | MA9 | Asymptomatic, Ward | White | 70's | Female | |
| | | MA10 | Mild, Ward | Other | 70's | Female | iabetes, HTN, A/COPD, Obesi |
| | | SEVY1 | ICU | Hispanic | 50's | Male | Diabetes, HTN |
| | | SEVY2 | Ward | Hispanic | 50's | Female | |
| | | SEVY3 | ICU | Hispanic | 30's | Female | Diabetes, HTN, CKD/ESRD |
| | | SEVY4 | Ward | Hispanic | 50's | Male | Diabetes, HTN |
| | | SEVY5 | Death | Hispanic | 20's | Male | Diabetes, HTN |
| | | SEVY6 | Death | Hispanic | 50's | Male | Diabetes, CKD/ESRD |
| | | SEVY7 | ICU | Hispanic | 30's | Male | Obesity |
| | | SEVY8 | ICU | Hispanic | 30's | Male | Obesity |
| | | SEVY9 | ICU | Hispanic | 40's | Male | HTN, Obesity |
| | | SEVY10 | ICU | Hispanic | 40's | Male | Diabetes, HTN |
| | | SEVY11 | ICU | Hispanic | 40's | Male | Diabetes, HTN, Obesity |
| | | SEVY12 | ICU | Native American | 40's | Male | HTN, Obesity |
| | | SEVY13 | ICU | Hispanic | 40's | Female | |
| | | SEVY14 | ICU | Hispanic | 50's | Male | |

| | | | | | | | |
|---|---|---|---|---|---|---|---|
| Severe | Young | SEVY15 | ICU/I | Hispanic | 20's | Male | A/COPD, Obesity |
| | | SEVY16 | ICU/I | Hispanic | 40's | Male | Obesity |
| | | SEVY17 | ICU/I | Hispanic | 50's | Male | |
| | | SEVY18 | ICU/I | White | 50's | Female | Diabetes, HTN, Obesity |
| | | SEVY19 | ICU/I | Hispanic | 50's | Male | Obesity |
| | | SEVY20 | ICU/I | Asian | 50's | Male | HTN |
| | | SEVY21 | ICU/I | Other | 50's | Female | Obesity |
| | | SEVY22 | Ward | Hispanic | 20's | Male | Obesity |
| | | SEVY23 | Ward | Unknown | 20's | Male | |
| | | SEVY24 | Ward | Hispanic | 30's | Female | Obesity |
| | | SEVY25 | Ward | Unknown | 30's | Female | |
| | | SEVY26 | Ward | Unknown | 30's | Male | |
| | | SEVY27 | Ward | White | 40's | Female | iabetes, HTN, A/COPD, Obesi |
| | | SEVY28 | Ward | White | 40's | Male | Diabetes, HTN |
| | | SEVY29 | Ward | Hispanic | 50's | Female | Diabetes, A/COPD, Obesity |
| | | SEVY30 | Ward | Hispanic | 50's | Male | HTN |
| | | SEVY31 | Ward | Other | 50's | Female | Obesity |
| | | SEVY32 | Ward | Unknown | 50's | Male | |
| | | SEVY33 | Ward | Hispanic | 50's | Female | Diabetes, HTN, CKD/ESRD |
| | | SEVY34 | Ward | Hispanic | 50's | Male | Obesity |
| | | SEVY35 | Ward | Hispanic | 50's | Male | HTN |
| | Aged | SEVA1 | Death | Hispanic | 60's | Male | HTN, Obesity |
| | | SEVA2 | Death | Hispanic | 70's | Male | Obesity |
| | | SEVA3 | Death | Hispanic | 90's | Female | HTN |
| | | SEVA4 | Death | Hispanic | 70's | Male | Diabetes, HTN, Obesity |
| | | SEVA5 | ICU | Asian | 80's | Female | HTN |
| | | SEVA6 | ICU | Hispanic | 60's | Female | Obesity |
| | | SEVA7 | ICU | White | 60's | Male | HTN, CVD |
| | | SEVA8 | ICU/I | Hispanic | 60's | Female | Diabetes, HTN, Obesity |
| | | SEVA9 | Ward | White | 60's | Male | |
| | | SEVA10 | Ward | Hispanic | 70's | Male | HTN, Obesity |
| | | SEVA11 | Ward | Hispanic | 60's | Female | Diabetes |
| | | SEVA12 | Ward | Hispanic | 60's | Male | |

| Antibody | Luminex | Flow | scRNAseq | Monocyte Phen | ICS | T cell peptide Stim |
|---|---|---|---|---|---|---|
| | Y | | | | | |
| | | Y | | | | |
| | Y | | | | | |
| | Y | | | | | |
| | Y | | | | | |
| | Y | | | | | |
| | | Y | | | | |
| | Y | | | | | |
| | Y | | | | | |
| | | Y | | | | |
| | Y | | | | | |
| | Y | | | | | |
| | Y | | | | | |
| | | Y | | | | |
| | | Y | | | | |
| | Y | | | | | |
| | | Y | | | | |
| | | Y | | | | |
| | | Y | | | | |
| | | Y | | | | |
| | | Y | | | | |
| | | Y | | | | |
| | | Y | | | | |
| | Y | | | | | |
| | Y | | | | | |
| | Y | | | | | |
| | Y | | | | | |
| | Y | | | | | |
| | Y | | | | | |
| | Y | | | | | |
| | Y | | | | | |
| | Y | | | | | |
| | Y | | | | | |
| | | | Y | | | Y |
| | | | Y | | | Y |
| | | | Y | | | Y |
| | | | Y | | | |
| | | Y | | | | |
| | | Y | | | | |
| | | Y | | | | |

| | | | | | | |
|---|---|---|---|---|---|---|
| | | Y | Y | | | |
| | | Y | | | Y | |
| | | Y | | | Y | |
| | | Y | Y | | | |
| | | Y | | | Y | |
| | | Y | | | Y | |
| | | Y | | | Y | |
| | | Y | Y | | | |
| | | Y | Y | | | |
| Y | Y | | | | | |
| Y | Y | | | | | |
| Y | Y | Y | | | | |
| Y | Y | | Y | | | |
| Y | Y | Y | Y | | | |
| Y | Y | Y | | | | |
| Y | | Y | Y | | | |
| Y | Y | Y | Y | | | |
| Y | Y | Y | | | | |
| Y | Y | | | | | |
| Y | Y | | | | | |
| Y | Y | | | | | |
| Y | Y | | | | | |
| Y | Y | | | | | |
| Y | | Y | | | | |
| Y | Y | Y | Y | | | |
| Y | Y | | Y | | | |
| Y | Y | Y | | | | |
| Y | Y | Y | Y | | | |
| Y | Y | | Y | | | |
| | | | | | | Y |
| | | | | | | Y |
| | | | | | | Y |
| | | | | | | Y |
| Y | Y | | | | | |
| Y | Y | | | | | |
| Y | | Y | | Y | Y | |
| Y | Y | | Y | | | Y |
| Y | Y | Y | | Y | Y | |
| Y | Y | Y | Y | Y | Y | Y |
| Y | Y | Y | | | | |
| Y | Y | Y | | | | |
| Y | Y | Y | | Y | Y | |
| Y | Y | Y | | Y | Y | |

| | | | | | | |
|---|---|---|---|---|---|---|
| Y | Y | Y | | | | |
| Y | Y | Y | Y | | | Y |
| Y | Y | Y | Y | | | Y |
| Y | Y | Y | | | | |
| Y | Y | Y | | Y | Y | |
| Y | Y | Y | | Y | Y | |
| Y | Y | Y | | Y | Y | |
| Y | Y | | | | | |
| Y | | Y | | | | Y |
| Y | Y | Y | | | | |
| Y | Y | Y | | | | |
| Y | Y | Y | | | | |
| Y | Y | Y | | Y | Y | |
| Y | Y | Y | | | | |
| Y | Y | Y | | Y | Y | |
| Y | Y | Y | | Y | Y | |
| Y | Y | Y | | | | |
| Y | Y | Y | | | | |
| Y | Y | Y | | | | |
| Y | Y | Y | | | | |
| Y | Y | Y | | Y | Y | |
| Y | Y | Y | | Y | Y | |
| Y | Y | | Y | | | |
| Y | Y | Y | | Y | Y | |
| Y | Y | Y | Y | | | |
| Y | Y | | | | | |
| Y | Y | Y | Y | | | |
| Y | Y | Y | | Y | Y | |
| Y | Y | Y | Y | | | |
| Y | Y | | | | | |
| Y | Y | | | | | |
| Y | Y | Y | | | | |
| Y | | Y | | | | |

| | Group | Total | %Female | Average Age (SEM) | % Hispanic |
|---|---|---|---|---|---|
| **Young** | Healthy Donors | 37 | 67.6 | 39.3 (1.6) | 16.2 |
| | Mild/Asymptomatic | 10 | 40.0 | 48.8 (3.3) | 50.0 |
| | Ward | 16 | 43.8 | 45.4(1.8) | 68.6 |
| | ICU | 10 | 20.0 | | |
| | ICU/I | 7 | 28.6 | | |
| | Fatality | 2 | 0.0 | | |
| **Aged** | Healthy Donors | 12 | 58.3 | 70.7 (3.1) | 0.0 |
| | Mild/Asymptomatic | 10 | 40.0 | 69.3 (1.4) | 50.0 |
| | Ward | 4 | 25.0 | 70.4 (2.6) | 75.0 |
| | ICU | 3 | 66.7 | | |
| | ICU/I | 1 | 100.0 | | |
| | Fatality | 4 | 25.0 | | |

| Average Age (SEM) | #Hispanic |
|---|---|
| 39.3243243243243 (1.66894171 | 6 |
| 48.8 [illegible] | 5 |
| 45.4 [illegible] | 24 |
| 70.75 [illegible] | 0 |
| 69.3 [illegible] | 5 |
| 70.4166666666667 [illegible] | 9 |

| % Comorbidities | Median DPS | SEM DPS |
|---|---|---|
| 2.7 | | |
| 80.0 | | |
| 77.1 | 11 | 0.57 |
| 0.0 | | |
| 90.0 | | |
| 83.3 | 10 | 0.85 |

| wout Comorbidities |
|---|
| 36 |
| 2 |
| 8 |
| 12 |
| 1 |
| 2 |

| |
|---|
| Diabetes |
| HTN |
| CVD |
| CAD |
| CKD/ESRD |
| Asthma/COPD |
| Obesity |

| |
|---|
| Young |
| Aged |

| |
|---|
| HC young |
| HC aged |
| Mild/Asympt young |
| Mild/Asympt aged |
| Severe young |
| Severe aged |

| |
|---|
| aged |
| HC |
| Mild/asymp |
| Severe |

| |
|---|
| Young |
| Mild/asymp |
| Severe |

| |
|---|
| Hypertension |
| Cardiovascular disease |
| Coronary Artery Disease |
| Chronic Kidney disease/ End stage renal disease |

| Median Age | % Female | Median DPS |
|---|---|---|
| 42.5 | 48.71794872 | 11 |
| 69 | 47.05882353 | 10 |

| Observed | Expected | Pvalue |
|---|---|---|
| 37 | 18.66666667 | 2.41447E-07 |
| 12 | 18.66666667 | |
| 10 | 18.66666667 | |
| 10 | 18.66666667 | |
| 31 | 18.66666667 | |
| 12 | 18.66666667 | |
| 112 | | |

| Observed | Expected | Pvalue |
|---|---|---|
| 12 | 11.33333333 | 0.889009765 |
| 10 | 11.33333333 | |
| 12 | 11.33333333 | |
| 34 | | |

| Observed | Expected | Pvalue |
|---|---|---|
| 10 | #REF! | |
| 31 | #REF! | |
| 41 | | |

Supplementary Table 2

*FindAllMarkers* function in Seurat was used to calculate the significant (q_value < 0.05) gen

| B cells | Plasmablasts | CD4 naïve | CD4 memory | Treg/prolif | CD8 naïve | CD8 memory |
|---|---|---|---|---|---|---|
| IGHM | JCHAIN | CCR7 | LTB | IL32 | CD8B | CCL5 |
| CD79A | IGHG1 | TSHZ2 | IL7R | DUSP4 | LINC02446 | GZMH |
| CD74 | IGHG3 | LEF1 | AP3M2 | STMN1 | NELL2 | NKG7 |
| HLA-DQA1 | IGHG2 | EEF1B2 | CRYBG1 | RGS1 | CD8A | CCL4 |
| HLA-DRA | MZB1 | PIK3IP1 | TNFRSF4 | TRAC | LEF1 | CD8A |
| CD83 | IGHA2 | RPS3A | MTRNR2L12 | CD52 | NDFIP1 | CST7 |
| HLA-DQB1 | IGHG4 | RPS12 | ANKRD12 | CYTOR | RPS5 | IFIT2 |
| MS4A1 | IGHGP | MAL | SESN3 | CTLA4 | CCR7 | GNLY |
| IGHD | ITM2C | IL7R | CDC14A | ARID5B | RPS12 | CD8B |
| TNFRSF13C | DERL3 | LTB | ARID5B | CORO1B | EEF1B2 | CTSW |
| HLA-DRB1 | PDIA4 | RPS13 | EML4 | CD3D | TRABD2A | CMC1 |
| IGKC | IGLL5 | RPL30 | RCAN3 | CD27 | LDHB | GZMA |
| BANK1 | CD38 | TCF7 | GPRIN3 | YWHAB | NUCB2 | TUBA4A |
| LINC00926 | TNFRSF17 | SARAF | ZFP36L2 | ICOS | RPS3A | CCL4L2 |
| HLA-DPB1 | PRDX4 | LDHB | ANK3 | SMC4 | RPL32 | IL32 |
| HLA-DPA1 | IGKV4-1 | LEPROTL1 | RORA | MIR4435-2H | PDE3B | GZMM |
| NFKBID | POU2AF1 | RCAN3 | BICDL1 | HELLS | RCAN3 | CD3D |
| IGLC2 | COBLL1 | RPL21 | MALAT1 | ARHGDIB | RPL10A | LYAR |
| CD37 | TXNDC5 | RPL32 | CD28 | STAM | RPL23 | TRGC2 |
| RALGPS2 | CD79A | NDFIP1 | SF1 | NPDC1 | RPL21 | KLRD1 |
| MKNK2 | SLAMF7 | RPS25 | TSHZ2 | CCDC167 | RPSA | FGFBP2 |
| CD79B | TNFRSF13B | RPS5 | AC058791.1 | MT1E | RPS21 | GZMK |
| MEF2C | HYOU1 | RPL13 | ICOS | PTTG1 | RPL13 | GZMB |
| HLA-DMB | SRPRB | RPL9 | GSPT1 | TIGIT | RPS6 | PMAIP1 |
| EZR | MYBL2 | RPS6 | ARHGAP15 | LINC01943 | RPL9 | CD3G |
| AFF3 | SPATS2 | RPL23 | TNFRSF25 | SKA2 | RPS13 | C12orf75 |
| IRF8 | SEL1L3 | RPL5 | SFXN1 | GMNN | RPL31 | RUNX3 |
| HLA-DMA | EAF2 | RPS23 | KLF2 | HMGB2 | RPS25 | IFNG |
| ADAM28 | FCRL5 | RPL34 | PABPC1 | FAM129A | RPS23 | DUSP2 |
| IGLC3 | MKI67 | RPS8 | SMCHD1 | LCK | RPL34 | LINC01871 |
| TCF4 | TCF4 | NOSIP | SARAF | TENT5C | RPL30 | HCST |
| SWAP70 | ISOC2 | TRABD2A | SPOCK2 | TMSB4X | RPL22 | CD3E |
| CD40 | SLC38A5 | RPS27 | PAG1 | CD5 | RPL12 | IFIT3 |
| BASP1 | CHST2 | RPL10A | DDX21 | LDHB | RPS18 | APOBEC3G |
| HLA-DRB5 | SEC24D | RPS14 | INPP4B | TRBC2 | RPS20 | OASL |
| REL | STT3A | RPL22 | DDX24 | CD28 | RPL5 | SYTL3 |
| CD22 | GGH | RPSA | ITK | JPT1 | RPS14 | XCL2 |
| MARCKS | TYMS | RPL36 | ZC3H12D | PCNA | RPS28 | TNF |
| SPIB | ST6GALNAC4 | ADTRP | PDE3B | SH2D1A | RPL14 | HOPX |
| COBLL1 | CHPF | RPS21 | FOXP1 | MZT2A | RPL7 | PPP2R5C |
| TCOF1 | RASGRP3 | RPL11 | SRSF7 | AES | RPS8 | LDHA |

| | | | | | | |
|---|---|---|---|---|---|---|
| FCER2 | ABCB9 | RPS27A | NDFIP1 | CD2 | APBA2 | CALM1 |
| BLK | RRM2 | EIF3E | KLF6 | PHF19 | RPL19 | TGFB1 |
| FCRL1 | CPNE5 | SNHG8 | TNFAIP3 | LAT | RPL27A | FYN |
| P2RX5 | GNG7 | RPS20 | CD6 | TTC39C | RPLP2 | PRF1 |
| PRDM2 | IGF1 | RPL31 | PRKCA | MIAT | RPL36 | ARPC5L |
| JUND | GMPPB | RPS28 | SERINC5 | BICDL1 | EEF1A1 | HLA-C |
| PAX5 | BHLHE41 | FHIT | MAL | PARP1 | RPL38 | KLRG1 |
| FAM129C | CEP128 | TPT1 | CAMK4 | MAF | RPL37 | METRNL |
| YBX3 | PHGDH | RPL35A | PASK | MT2A | RPL35A | PIK3R1 |
| BCL11A | GAS6 | RPL10 | BCL2 | CRIP1 | RPL37A | AKNA |
| GRASP | CLIC4 | RPS29 | BTG1 | ITGB1 | RPS29 | TRBC2 |
| TSPAN33 | GAB1 | RPS15A | AQP3 | SOD1 | RPL18 | JMJD6 |
| PHACTR1 | PCLAF | RPL38 | GATA3 | AQP3 | RPL10 | TRBC1 |
| ATF7IP | NT5DC2 | RPL24 | FXYD5 | CDC25B | RPS27A | BHLHE40 |
| HERPUD1 | SLC17A9 | SOCS3 | FUS | PHACTR2 | RPL23A | PAXX |
| LAPTM5 | LINC01480 | RPS18 | LEPROTL1 | FAS | RPS2 | CREM |
| TRAF4 | TRAM2 | GIMAP7 | BCL11B | CD3G | RPL28 | AUTS2 |

es for each cluster

| NK | NKT | Monocytes C | Monocytes C | DC | Progenitor Cl | Erythroid |
|---|---|---|---|---|---|---|
| GNLY | SYNE1 | S100A8 | LST1 | HLA-DRA | SOX4 | HBB |
| GZMB | SYNE2 | S100A9 | AIF1 | HLA-DPA1 | TCF4 | HBA2 |
| PTGDS | RNF213 | LYZ | FCGR3A | HLA-DRB1 | ITM2C | HBA1 |
| NKG7 | MACF1 | CXCL8 | IFITM3 | HLA-DPB1 | BCL11A | ALAS2 |
| SPON2 | UTRN | S100A12 | COTL1 | CST3 | PLD4 | CA1 |
| CCL3 | AKNA | VCAN | CDKN1C | CD74 | SERPINF1 | AHSP |
| FGFBP2 | ZEB2 | FCN1 | CST3 | HLA-DQA1 | AVP | HBD |
| PRF1 | MTRNR2L12 | CTSS | FCER1G | HLA-DQB1 | PRSS57 | HBM |
| GZMA | PRF1 | NAMPT | SERPINA1 | HLA-DRB5 | TPM2 | SNCA |
| CCL4 | MT-ATP6 | AC020656.1 | FTL | GPR183 | SPINK2 | SLC25A37 |
| CST7 | MT-ND5 | CST3 | PSAP | HLA-DMA | STMN1 | UBB |
| KLRD1 | MT-CO2 | EREG | S100A11 | LYZ | AC084033.3 | SLC25A39 |
| KLRB1 | MT-CO1 | DUSP6 | MT2A | CLEC10A | GNA15 | BLVRB |
| CTSW | MALAT1 | MAFB | SAT1 | FCER1A | MZB1 | BPGM |
| KLRF1 | MT-ND1 | G0S2 | CFD | CEBPD | LILRA4 | BNIP3L |
| CD247 | MT-ND2 | CEBPD | APOBEC3A | CPVL | SPINT2 | GLRX5 |
| CLIC3 | MT-CO3 | AIF1 | MS4A7 | ANXA2 | DDAH2 | FAM210B |
| CD7 | MT-ND4 | CD14 | SPI1 | GSTP1 | FAM30A | YBX3 |
| FCGR3A | MT-ND3 | IFI27 | CTSS | CTSZ | SCT | SELENBP1 |
| HOPX | MT-CYB | IL1B | C5AR1 | KLF4 | SMPD3 | GYPC |
| IL2RB | KLRD1 | THBS1 | TYROBP | HLA-DMB | GATA2 | SLC4A1 |
| CCL5 | PRRC2C | SERPINA1 | SMIM25 | LGALS2 | IL3RA | DCAF12 |
| MYOM2 | PTPRC | GRN | LILRB2 | PHACTR1 | EGFL7 | FKBP8 |
| GZMM | NEAT1 | PSAP | LYN | CCDC88A | PTMS | PRDX2 |
| EFHD2 | FLNA | S100A11 | TIMP1 | EREG | TSC22D1 | BCL2L1 |
| GNG2 | SYTL3 | S100A6 | FTH1 | CTSH | SMIM24 | HEMGN |
| ABHD17A | RBM39 | TYROBP | WARS | GRN | DERL3 | IFIT1B |
| XCL2 | CCL5 | TYMP | HLA-DPA1 | TYMP | RASD1 | MPP1 |
| S1PR5 | NKTR | IFITM3 | CD68 | SPI1 | TSPAN13 | BSG |
| TRDC | ITGAL | KLF4 | LRRC25 | AC020656.1 | AC119428.2 | STRADB |
| IGFBP7 | SYTL2 | FTL | ASAH1 | FCGRT | CRHBP | ADIPOR1 |
| HCST | SRRM2 | PLAUR | MARCKS | IL1B | CYTL1 | HAGH |
| GZMH | MYO1F | NEAT1 | HES4 | CAPG | BCL7A | LGALS3 |
| MATK | HNRNPH1 | SPI1 | NPC2 | AP1S2 | LAPTM4B | MKRN1 |
| CMC1 | ZAP70 | FCER1G | VSIR | CD1C | TFPI | TRIM58 |
| RUNX3 | AKAP13 | LST1 | TYMP | C1orf162 | LGMN | GABARAPL2 |
| IFNG | RUNX3 | PLBD1 | PECAM1 | ANXA5 | CD34 | HBQ1 |
| ADGRG1 | IRF1 | CEBPB | G0S2 | HBEGF | FCER1A | GMPR |
| APMAP | TCF25 | CLEC7A | MAFB | NR4A1 | SMIM3 | GUK1 |
| OTULIN | ADGRG1 | C5AR1 | FCN1 | PLAUR | MAP1A | KRT1 |
| CCL4L2 | MYBL1 | CYBB | S100A4 | RAB31 | FHL1 | NCOA4 |

| | | | | | | |
|---|---|---|---|---|---|---|
| METRNL | PPP2R5C | SAT1 | CYBB | VEGFA | CBFA2T3 | DMTN |
| MAFF | CEP78 | MNDA | RHOC | AIF1 | ALDH1A1 | EPB42 |
| ARL4C | FYN | CSTA | S100A6 | LST1 | CLEC11A | FECH |
| ARHGAP9 | MT-ND4L | MARCKS | CEBPB | CFP | MYL6B | TMCC2 |
| DSTN | SLFN12L | SLC11A1 | IFITM2 | CDKN1A | RAB13 | FBXO7 |
| BHLHE40 | FUS | APLP2 | FGL2 | CSF2RA | NPR3 | RNF10 |
| IFITM2 | SON | CSF3R | TCF7L2 | AP2S1 | PTCRA | MAP2K3 |
| PMAIP1 | GNPTAB | LGALS1 | HLA-DRB5 | RNF130 | MSRB3 | OSBP2 |
| ITGB2 | STAT4 | CTSD | HLA-DRA | ALDH2 | LINC02573 | HMBS |
| GRASP | PCSK7 | COTL1 | BRI3 | CSTA | IL18 | PDZK1IP1 |
| SYNE1 | GZMB | TSPO | TNFRSF1B | BID | ZNRF1 | LYL1 |
| JAK1 | SPON2 | S100A10 | NEAT1 | LY86 | MDK | AC092490.1 |
| IER2 | PRPF4B | SGK1 | LILRA5 | PYCARD | AL157895.1 | GSPT1 |
| PFN1 | CEMIP2 | MS4A6A | HMOX1 | JAML | MYB | MXI1 |
| AREG | DYNC1H1 | S100A4 | PILRA | PLSCR1 | SHD | EIF2AK1 |
| GNPTAB | AKAP9 | TKT | IFI30 | LMO2 | CIB2 | IFI27 |
| LAIR2 | APOL6 | BRI3 | ANXA5 | IFI30 | NREP | GYPB |

**Platelets**
PPBP
PF4
NRGN
GNG11
CAVIN2
TUBB1
MYL9
CLU
RGS18
HIST1H2AC
TREML1
SPARC
GP9
ITGA2B
PRKAR2B
ACRBP
CMTM5
LIMS1
F13A1
TLN1
MPIG6B
NCOA4
CTSA
VCL
FERMT3
BEX3
CD9
2-Mar
OAZ1
RGS10
PGRMC1
ARPC1B
OST4
YWHAH
TPM1
TPM4
TSC22D1
TAGLN2
PDLIM1
HIST1H3H
PTCRA

CLEC1B
RUFY1
TMEM40
MTURN
CALM3
PARVB
ILK
TIMP1
C2orf88
SNCA
LAMTOR1
CD151
CTTN
MPP1
GRAP2
PTGS1
SH3BGRL3
